# Racism and health and wellbeing among children and youth - an updated systematic review and meta-analysis

**DOI:** 10.1101/2024.07.18.24310663

**Authors:** Naomi Priest, Chiao Kee Lim, Kate Doery, Jourdyn A. Lawrence, Georgia Zoumboulis, Gabriella King, Dewan Lamisa, Fan He, Rushani Wijesuriya, Camila M. Mateo, Shiau Chong, Mandy Truong, Ryan Perry, Paula Toko King, Natalie Paki Paki, Corey Joseph, Dot Pagram, Roshini Balasooriya Lekamge, Gosia Mikolajczak, Emily Darnett, Brigid Trenerry, Shloka Jha, Joan Gakii Masunga, Yin Paradies, Yvonne Kelly, Saffron Karlsen, Shuaijun Guo

## Abstract

**Background:** Evidence of racism’s health harms among children and youth is rapidly increasing, though attention to impacts on physical health and biomarker outcomes is more emergent. We performed a systematic review of recent publications to examine the association between racism and health among children and youth, with a meta-analysis of the specific relationships between racism and physical health and biomarkers.

**Methods:** We conducted a systematic literature search using four databases: Medline, PsycINFO, PubMed, and ERIC. Four inclusion criteria were used to identify eligible studies: (1) exposure was experiences of racism, (2) outcome was health and wellbeing, (3) quantitative methods were used to estimate the association between racism and health outcomes, and (4) the effect size of associations between racism and health and wellbeing was reported for participants aged 0-24 years. Correlation coefficients were used to report the pooled effect size for each outcome indicator.

**Results:** There were 463 eligible studies included in the screening process, with 42 studies focusing on physical health or biomarker outcomes. Random-effects meta-analysis found minimal to moderate positive associations between racism and C-reactive protein, Interleukin 6, body mass index (BMI), obesity, systolic blood pressure, salivary cortisol, asthma, and somatic symptoms. There were marginal positive associations between racism and Tumour Necrosis Factor-α, cortisol collected via saliva, urine and hair, BMI-z score, and diastolic blood pressure, with imprecise estimates and wide confidence intervals.

**Conclusions:** Racism is associated with negative physical health and biomarker outcomes that relate to multiple physiological systems and biological processes in childhood and adolescence. This has implications for health and wellbeing during childhood and adolescence and future chronic disease risk. Collective and structural changes to eliminate racism and create a healthy and equitable future for all children and youth are urgently required.

**HIGHLIGHTS:**

- Racism impacts foundations for optimal health and development for children, youth and their caregivers.
- Evidence shows racism is associated with inflammation, cortisol, weight status, blood pressure, asthma and somatic symptoms in children and youth.
- Future research needs to prioritise systemic racism and its impact on child and youth health.
- Collective and structural changes are urgently needed to eliminate racism.

## INTRODUCTION

Racism is a pervasive and persistent social, political, and human rights issue (Cramer, 2020; Parter et al., 2023) and threat to public health globally (Devakumar et al., 2020). Racism is an organised system of oppression that creates race by categorising and ranking social groups and devalues, disempowers and differentially allocates desirable societal opportunities and resources to groups considered inferior (Bonilla-Silva, 1996; Williams, 2004). The relegation of racialised groups to inferior status and treatment, and intentional and unintentional unfair treatment and oppression of people from racialised groups are all part of the system of racism (Braveman et al., 2022). Racism is a fundamental cause and powerful determinant of health and health inequities that harms health via multiple complex pathways operating throughout the life course and across generations (Braveman et al., 2022; Williams & Mohammed, 2013). Children and youth are particularly vulnerable to racism’s harms (Gee et al., 2012; Trent et al., 2019), with childhood and adolescence recognised as sensitive periods during which social and environmental exposures interact with and shape biological processes to influence lifelong health (Patton et al., 2016; Shonkoff et al., 2021).

Socio-ecological models that highlight the ecological systems in which children grow and develop are foundational to our understanding of child and youth health and development (Shonkoff et al., 2012). Integrating these ecological models of child development (Shonkoff et al., 2012) with models of racism and health (Williams & Mohammed, 2013) emphasises the foundational role that systemic racism plays in driving health and health inequities for children and youth from racialised groups and the ways in which systemic racism impacts foundations for optimal development for children, youth and their caregivers (Slopen & Heard-Garris, 2022). Systemic and structural racism are forms of racism that are widely and deeply entrenched within and across societal systems, laws, policies and practices written and unwritten, and values, beliefs and attitudes explicit and implicit (Bailey et al., 2017; Braveman et al., 2022; Gee & Hicken, 2021; Michaels et al., 2023). Reflecting ongoing and historical injustices, systemic and structural racism create, perpetuate, and reinforce unjust and unfair treatment of people from racialised groups via interconnected mutually reinforcing inequitable systems (Braveman et al., 2022; Michaels et al., 2023). Institutional racism is often used interchangeably with systemic or structural racism, though also refers more specifically to racism within particular institutions rather than the interconnected nature of racism across systems of institutions (Braveman et al., 2022; Jones, 2000; Michaels et al., 2023; Williams et al., 2019).

Systemic racism harms the health of children and youth via multiple direct and indirect pathways that impact them, their families and their communities and create physical, social and economic conditions that threaten their health, development and wellbeing (Priest et al., 2013; Shonkoff et al., 2021). Systemic racism limits access to opportunities such as education, employment, financial security, housing and neighbourhood safety resulting in socioeconomic inequities (Priest et al., 2013; Shonkoff et al., 2021). Directly, and through socioeconomic pathways, racism can influence health behaviours such as physical activity, diet, smoking, alcohol and drug use, and shapes exposure to physical, chemical and psychosocial stressors that can influence health through behavioural, psychological, and biological pathways (Priest et al., 2013; Shonkoff et al., 2021). Through each of these pathways, structural racism influences the social and physical environment surrounding children and youth and impacts foundations for optimal caregiving and child and youth development (Slopen & Heard-Garris, 2022). Interpersonal racism, the everyday racism that occurs between individuals, is a further enactment of structural racism that is itself an important stressor contributing to health and health inequities for children and youth (Nazroo et al., 2020; Priest et al., 2023; Priest et al., 2013; Shonkoff et al., 2021). This includes direct interpersonal racism where children and youth themselves are targets and vicarious experiences where children and youth witness racism directed at others (Heard-Garris et al., 2017; Priest et al., 2013). Caregiver experiences of interpersonal racism, whether they are witnessed by children or not, can also have harmful effects on child and youth health including via impacts on caregiver mental and physical health, parenting, and the accumulation of health-harming contexts and experiences (Heard-Garris et al., 2017; Priest et al., 2013; Slopen & Heard-Garris, 2022). A further form of racism that is driven by, and reinforces systemic racism, is internalised racism; defined as the internalisation of inferior status and treatment by racialised groups (Williams et al., 2019). Reflecting systemic racism not individual capabilities or supposed pathologies, internalised racism further limits caregivers’ and children’s access to health supporting resources and opportunities (Williams et al., 2019).

Echoing what First Nations peoples and people of colour have been saying for centuries, recent international systematic reviews have empirically documented the harmful impacts of racism on child and youth health; most of these reviews narratively synthesised findings (Heard-Garris et al., 2017; Priest et al., 2013; Uink et al., 2022), with one meta-analysis (Benner et al., 2018). One further review examined the longitudinal effects of racism exposure in childhood on health, however, findings were difficult to interpret due to lack of reporting of age of outcomes (Cave et al., 2020). Findings from these reviews highlight that the predominant focus of existing evidence on interpersonal racism often lacks grounding in conceptual frameworks of racism that highlight its systemic nature and the pathways and processes by which systemic racism drives poor health, and for Indigenous Peoples that recognise and acknowledge the profoundly interlinked nature of racism, settler-colonialism and other forms of oppression (Heard-Garris et al., 2017; Priest et al., 2013; Uink et al., 2022). Negative impacts of interpersonal racism on youth mental health outcomes are most commonly reported across the reviews (Benner et al., 2018; Heard-Garris et al., 2017; Priest et al., 2013; Uink et al., 2022), with a meta-analysis finding the strongest effects for depression (*r*=0.26) and internalising symptoms (*r*=0.26). Negative effects of interpersonal racism on youth risky health behaviours were the next most common finding (Benner et al., 2018; Cave et al., 2020; Priest et al., 2013), with a small to moderate effect size (*r*=0.20) (Benner et al., 2018). Evidence of negative effects of interpersonal racism on mental health for preschool and primary/elementary school-aged children have also been reported by the reviews that considered outcomes for pre-adolescents (Heard-Garris et al., 2017; Priest et al., 2013). Across the reviews, physical health outcomes were far less commonly reported and with mixed findings. Priest et al (Priest et al., 2013) reported only 15 of 461 examined associations were related to physical health across diverse outcomes such as blood pressure, childhood illnesses, insulin resistance and obesity; 67% of these showed no evidence of association. Heard-Garris et al (Heard-Garris et al., 2017) identified five reported associations between vicarious racism and physical health (aside from birthweight and preterm birth), with evidence of associations in one study for cortisol reactivity, body mass index (BMI), weight for age and mixed findings for general childhood illness.

Notwithstanding the importance of documenting mental health and behavioural impacts of racism, key knowledge gaps clearly remain regarding the relationship between systemic racism and child and youth physical health and the biological mechanisms by which racism becomes embedded and embodied in early life. Such knowledge of biological mechanisms is essential to evaluating causality and necessary for understanding the pathways, material, psychosocial, behavioural and biological, by which systemic racism becomes embodied and biologically expressed in individual’s health and population-level health inequities (Krieger, 2012; Krieger, 2014). This is particularly crucial in childhood and adolescence where biological systems are especially sensitive to social and environmental influences with resulting structural changes and physiological disruptions having lifelong health impacts (Shonkoff et al., 2021). Critically, such knowledge is not needed to establish that racism is bad; racism is by definition unfair, and is a breach of human rights (Krieger, 2000; Krieger, 2003). Rather greater understanding of such mechanisms is required to inform accurate explanations for health inequities and effective interventions to address them, and to deter those that are false and harmful (Krieger, 2000).

The overarching aim of this review is to examine recent evidence on the extent to which racism is associated with health and wellbeing among children and youth, updating the previous systematic review (Priest et al., 2013). Informed by key gaps in the literature as reviewed above, we focused the main review and meta-analysis on physical health and biomarker outcomes. Characteristics of studies identified in the initial search are outlined to provide context for the more targeted review and meta-analysis on physical health and biomarker outcomes.

## METHODS

To ensure the reporting quality of this review, we followed the previously published protocol (Priest et al., 2021), the Preferred Reporting Items for Systematic Reviews and Meta-Analysis (PRISMA) 2020 checklist (Matthew et al., 2021), and the Meta-analysis of Observational Studies in Epidemiology (MOOSE) guidelines (Stroup et al., 2000).

### Literature search

We conducted a literature search using four databases: MEDLINE (Ovid), PsycINFO, PubMed, and ERIC (EBSCO) for studies published up until 16 December 2022. These databases span biomedical and health sciences, psychological, behavioural and social sciences and education peer-reviewed journals, books and book chapters, and reports (See Supplementary File 1 for full details of the search strategy). In brief, three types of search terms were used: (1) exposure-related terms: e.g., ‘*racism*’ OR ‘*race*’; (2) outcome-related terms: e.g., ‘*distress*’ OR ‘*biomarker*’; and (3) age-related terms: e.g., ‘*child*’ OR ‘*adolescence*’.

### Eligibility criteria

Studies had to fulfil the following criteria to be included: (1) participants or those whose experiences were being reported were children or youth aged 0 to 24 years old. Both self and proxy reports (e.g., caregivers, teachers) of experiences of racism were included; (2) the exposure was experiences of racism, including structural, institutional and interpersonal experiences of racism, including direct, vicarious and internalised racism; (3) the outcome focused on health and wellbeing, which included: (a) pregnancy and birth outcomes, (b) general health and wellbeing, (c) physical health, (d) negative mental health, (e) positive mental health, (f) health behaviours and sleep, (g) healthcare utilisation, healthcare costs, satisfaction with the healthcare system, and (h) biological markers; and (4) the study used quantitative methods reporting an estimate of the association between racism and health and wellbeing.

Studies were excluded if: (1) they were published before 2012, to build on the results of the previous systematic review (Priest et al., 2013); (2) the full-text papers were not available; (3) they used qualitative methods or did not report associations between racism and health; (4) they were published in a language other than English; or (5) they did not attribute experiences of discrimination to race or ethnicity (i.e. considered discrimination based on other demographic characteristics such as gender and disability).

### Selection of studies

Search results were initially imported into EndNote 20, which were then exported into Covidence (Babineau, 2014), a web-based systematic review software. Duplicates were removed before title and abstract screening. Next, 15 reviewers (SG, KD, JL, MT, PTK, NPP, NP, YP, RP, GK, ED, BT, GZ, RB, and GM) independently screened all studies based on the title and abstract. Full-text papers of the remaining studies were then obtained and reviewed independently by 17 reviewers (SG, DL, NP, RB, JL, GK, GZ, MT, PTK, NPP, ED, DP, FH, KD, RP, BT, and SJ). At each step of the screening process, discrepancies between reviewers were resolved by discussion with a third reviewer or by consensus (SG, KD, JL, NP). The study selection process is outlined in Figure 1.

**Figure 1.**
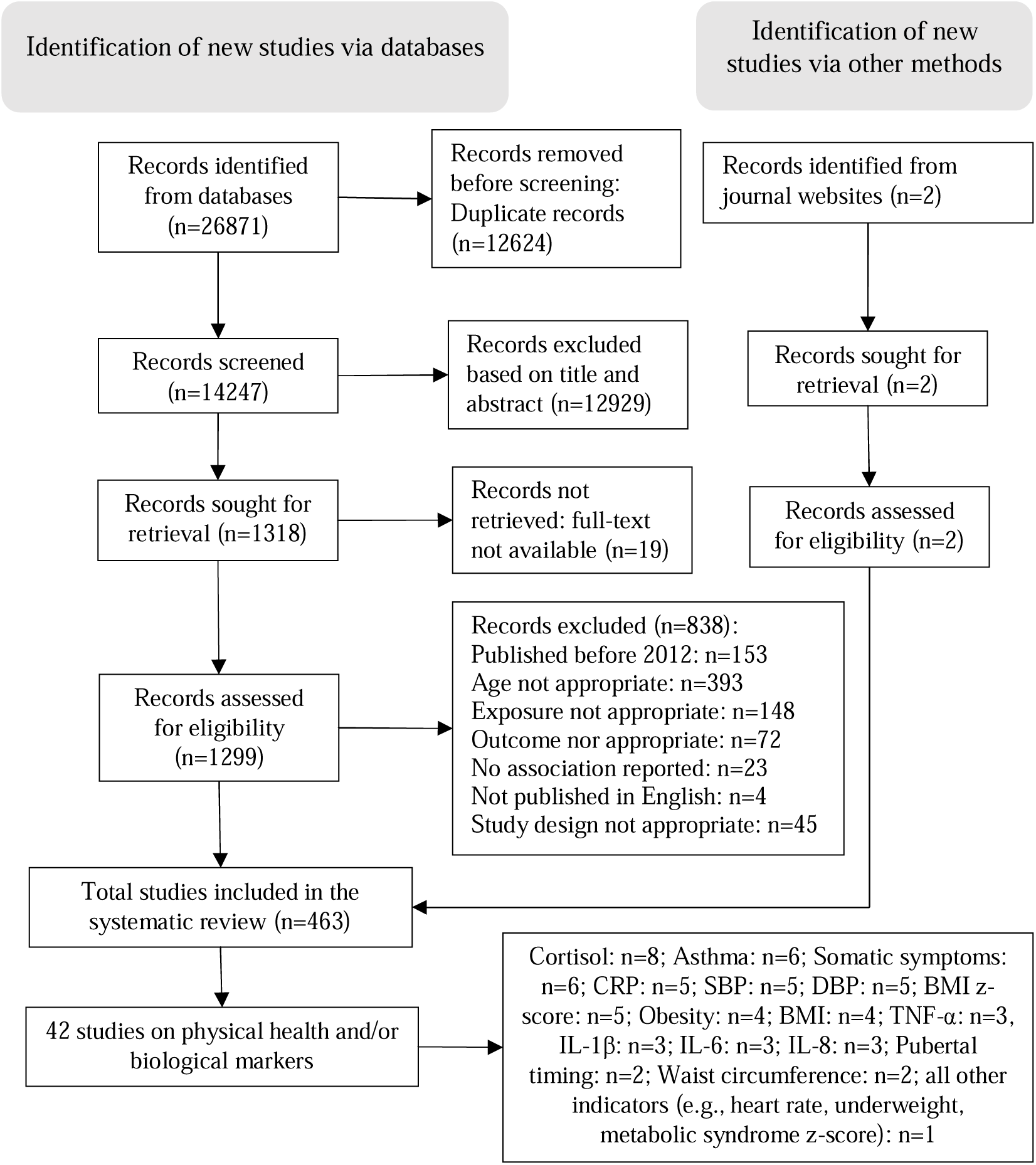
Flow chart of search and selection process according to the 2020 Preferred Reporting Items for Systematic Reviews and Meta-Analyses flow diagram

### Data extraction and analysis

Data from identified studies were extracted into Airtable (Howie Liu et al., 2012) in two steps.

Step one mapped characteristics of studies on racism and child health identified since the previous review (Priest et al., 2013) to provide context for the focused review and meta-analysis (step two). Data were extracted from all included studies by 15 reviewers (SG, FH, NPP, PTK, DP, RB, LD, JL, GM, CJ, MT, GK, GZ, CM, CKL), with one reviewer (CKL) randomly checking 5% of the extracted data (Vale et al., 2004). Extracted data included information on study characteristics (e.g., first author, published year, country of location), demographics of participants (e.g., age, gender, race), exposure type (e.g., direct self-report, proxy parent-report, proxy teacher-report), outcome type (e.g., mental health, health behaviours and sleep, biological markers), the definition of racism if it was specified, and the underlying theory or framework if any was used. Study characteristics were narratively synthesised following the approach of the previous review (Priest et al., 2013).

Step two focused on data extraction from studies that reported physical health or biomarker outcomes for the main review and meta-analysis. Information on exposure, (e.g., indicator, instrument, psychometric properties), outcome (e.g., indicator, instrument, psychometric properties), and associations between racism and child and youth health and wellbeing (e.g., sample size, statistical methods, effect size) were extracted by three reviewers (SG, CKL, SC), with one reviewer (CKL) randomly checking 20% of the extracted data (Lawrence et al., 2022). Discrepancies were resolved by consensus or discussion with third reviewers. We also critically appraised those studies included in the meta-analysis. Non-randomised studies (i.e., cross-sectional, case-control, cohort) were assessed using the Newcastle-Ottawa Scale (Wells et al., 2019). Experimental studies such as randomised controlled trials were assessed using the Joanna Briggs Institute Critical Appraisal quality of these studies, with one reviewer (CKL) randomly checking 20% of the extracted data. Discrepancies were resolved by consensus or discussion with third reviewers. The quality ratings for each study are provided in Supplementary File 2.

Correlation (*r*) coefficients were the most commonly reported measure of associations and were used as the principal measure of effect size for the meta-analysis. Standardised mean differences (i.e., standardised betas; *β*) in unadjusted regression models are considered equal to correlation coefficients (Bowman, 2012). We used the following formula to transform adjusted standardised mean differences to correlation coefficients, where *λ* is an indicator variable that equals 1 when *β* is positive and 0 when *β* is negative (Peterson & Brown, 2005):

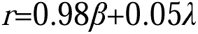

When unstandardised mean differences (*B*) were presented in the papers, they were converted to standardised mean differences before incorporation into the analysis using the following formula (Peterson & Brown, 2005):

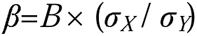

where σ*_X_* is the standard deviation of the independent variable and σ*_Y_* is standard deviation of the dependent variable. When risk ratios (RRs) were reported, we transformed them into odds ratios (ORs) using the following formula (Wang, 2013):

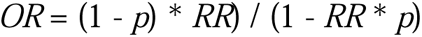

where *p* is the risk in the reference group. ORs were then transformed to correlation coefficients using the following formulas (Borenstein et al., 2009):

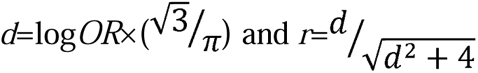

where *d* is the standardised mean difference and *r* is the correlation coefficient.

Meta-analyses were conducted using the ‘*meta*’ package available in Stata 18.0 (StataCorp, 2023). In keeping with previous meta-analyses (Lawrence et al., 2022), when the same evidence was presented in multiple papers, we extracted data only from the paper that reported the most detailed information on effect size. Where the level of detail was the same, the earliest publication was included in the meta-analysis. When a paper reported multiple associations between racism and health outcomes for the same participants, we used effect sizes at the level of individual outcomes (i.e., for each outcome indicator such as cortisol, BMI, and asthma) to avoid a violation of analytical independence assumptions and calculated a pooled effect size when there were at least three studies for the same association (van Daalen et al., 2022). Forest plots are presented to illustrate study-specific and overall effect sizes and 95% confidence intervals (CI) by each outcome indicator.

We did not produce an overall effect size for the relationship between racism and physical health and biomarkers, and instead relied on each specific outcome indicator (e.g., C-reactive protein (CRP), cortisol, BMI) to generate a specific effect size. This was to limit the effect of measurement bias in the final findings that may arise due to differences in scales and to ensure the findings were meaningful to interpret.

We extracted both minimally and maximally adjusted estimates of effect sizes where possible. Minimally adjusted estimates included data from the least adjusted model reported or correlations between racism and health outcomes. Maximally adjusted estimates included data from the most adjusted model reported with all covariates included. However, if only one estimate was available, it was used as both the minimally and maximally adjusted estimate, although efforts were made to contact study authors for additional information.

In the main text, we used the minimally adjusted estimates for meta-analyses. Due to the expected heterogeneity between studies, random effects models were performed, assuming a normal distribution of effect sizes allowing for variations in the effect size across studies, where factors beyond sampling variation may influence the association (Borenstein et al., 2010). We conducted Cochran’s Q test and generated an I^2^ statistic as a percentage of variability, with 75%, 50% and 25% indicating high, moderate, and low heterogeneity (Melsen et al., 2014). Funnel plots and Egger’s tests were used to examine the possibility of publication bias (Sedgwick & Marston, 2015). We also conducted sensitivity analyses to check the robustness of our findings, which estimated the effect sizes using the maximally adjusted estimates reported in eligible articles.

## RESULTS

The initial search identified 27,962 studies. After the removal of duplicates and initial title/abstract screening, 1,320 full-text papers were identified for further assessment. After applying the eligibility criteria, 463 studies were included in the initial context mapping, including 42 studies that had physical health or biomarker outcomes and were subsequently included in the meta-analysis (see Figure 1). Our checks showed consistency for 98% (step one) and 99% (step two) of extracted records.

### Step one: Narrative synthesis results (n=463)

Key characteristics of all included studies (n=463) are presented in Table 1 (see Supplementary File 3 for details). The pace of publishing was roughly steady in the last ten years, with almost one-third of papers (n=149, 32%) published since 2021. Most studies used a cross-sectional study design (n=255, 55%), collected data from convenience (non-representative) samples (n=371, 80%), and had a sample size of 200 or more (n=347, 75%). Most studies were conducted in the USA (n=381, 82%) and focused on Black participants (n=272, 59%). Overall, these included studies captured experiences of racism at different life stages (newborn: n=31, preschool: n=17, primary school: n=118, secondary school: n=352, post-school: n=110).

**Table 1.** Characteristics of included studies that reported racism and child health.

|  | n (%) |  |
| --- | --- | --- |
|  | All included studies<br>(n=463) | Studies that had physical health or<br>biomarker outcomes (n=42) |
| **Year of publication** |  |  |
| 2012-2015 | 123 (26.6) | 13 (31.0) |
| 2016-2020 | 191 (41.3) | 18 (42.9) |
| 2021-2023 | 149 (32.2) | 11 (26.2) |
| **Publication type** |  |  |
| Academic journal | 427 (92.2) | 40 (95.2) |
| Dissertation/report/book chapter | 36 (7.8) | 2 (4.8) |
| **Study design** |  |  |
| Case-control | 3 (0.6) | 2 (4.8) |
| Cross-sectional | 255 (55.1) | 17 (40.5) |
| Longitudinal | 203 (43.8) | 22 (52.4) |
| Experimental | 2 (0.4) | 1 (2.4) |
| **Sampling procedure** |  |  |
| Convenience | 371 (80.1) | 35 (83.3) |
| Representative | 92 (19.9) | 7 (16.7) |
| **Sample size** |  |  |
| n<100 | 29 (6.3) | 6 (14.3) |
| 100≤n<200 | 86 (18.6) | 9 (21.4) |
| 200≤n<500 | 138 (29.9) | 10 (23.8) |
| 500≤n<1000 | 103 (22.3) | 5 (11.9) |
| n≥1000 | 106 (22.9) | 12 (28.6) |
| **Region of study** |  |  |
| Australia/New Zealand | 18 (3.9) | 6 (14.3) |
| South America | 4 (0.9) | 0 |
| Canada | 7 (1.5) | 0 |
| Europe/UK | 40 (8.6) | 3 (7.1) |
| Asia | 10 (2.2) | 0 |
| USA | 381 (82.3) | 33 (78.6) |
| Two or more regions | 3 (0.6) | 0 |
| **Study population characteristics** |  |  |
| *Age group\** |  |  |
| Newborns/infants (0-2 years) | 31 (6.7) | 2 (4.8) |
| Preschool (3-5 years) | 17 (3.7) | 7 (16.7) |
| Primary school (6-11 years) | 118 (25.5) | 15 (35.7) |
| Secondary school (12-18 years) | 352 (76.0) | 25 (59.5) |
| Post-school (19-24 years) | 110 (23.8) | 25 (59.5) |
| *Child's gender\** |  |  |
| Male and female | 416 (89.8) | 37 (88.1) |
| Female only | 18 (3.9) | 1 (2.4) |
| Male only | 14 (3.0) | 1 (2.4) |
| Other | 15 (3.2) | 1 (2.4) |
| *Ethnic/racial group\** |  |  |
| Anglo-Celtic/White | 93 (20.1) | 7 (16.7) |
| European | 32 (6.9) | 4 (9.5) |
| Latino/a | 191 (41.3) | 21 (50.0) |
| African American/Black | 272 (58.7) | 23 (54.8) |
| Asian | 116 (25.1) | 10 (23.8) |
| Indigenous | 66 (14.3) | 7 (16.7) |
| African | 7 (1.5) | 0 |
| Middle Eastern | 33 (7.1) | 3 (7.1) |
| Other | 113 (24.4) | 13 (31.0) |
| *Refugee/immigrant status\** |  |  |
| Refugees | 5 (1.1) | 0 |
| Native born | 148 (32.0) | 18 (42.9) |
|  | All included studies<br>(n=463) | Studies that had physical health or<br>biomarker outcomes (n=42) |
| Born overseas | 151 (32.6) | 18 (42.9) |
| *Place of residence\** |  |  |
| Urban | 201 (43.4) | 19 (45.2) |
| Rural | 71 (15.3) | 5 (11.9) |
| Remote | 12 (2.6) | 3 (7.1) |
| *Racism is defined* |  |  |
| No | 285 (61.6) | 24 (57.1) |
| Yes | 178 (38.4) | 18 (42.9) |
| *Conceptual model is used* |  |  |
| No | 193 (41.7) | 24 (57.1) |
| Yes | 270 (58.3) | 18 (42.9) |
| *Type of racial discrimination\** |  |  |
| Direct (self-report) | 430 (92.9) | 38 (90.5) |
| Direct (parent/teacher-report) | 41 (8.9) | 9 (21.4) |
| Vicarious | 21 (4.5) | 2 (4.8) |
| Institutional/Structural | 28 (6.0) | 1 (2.4) |
| Internalised | 4 (0.9) | 1 (2.4) |
| *Type of health outcome\** |  |  |
| Pregnancy and birth outcomes | 21 (4.5) | 0 |
| General health and wellbeing, life<br>satisfaction and quality of life | 33 (7.1) | 5 (11.9) |
| Physical health | 33 (7.1) | 33 (78.6) |
| Negative mental health | 309 (66.7) | 13 (31.0) |
| Positive mental health | 82 (17.7) | 2 (4.8) |
| Health behaviours and sleep | 126 (27.2) | 7 (16.7) |
| Healthcare utilisation, healthcare costs,<br>and satisfaction | 9 (1.9) | 0 |
| Biological markers | 16 (3.5) | 16 (38.1) |

Note: \* Categories are not mutually exclusive in relation to the unit of analysis (i.e., studies).

Fewer than half of the included studies (n=178, 38%) provided an explicit definition of racism in the context of a paper prior to the methods section. Among those, 48 studies defined racism at the structural level and 22 at the institutional level (see Supplementary File 3 for details). Most studies defined racism at the interpersonal level, with direct racial discrimination defined in 141 studies and vicarious racial discrimination defined in five studies. Seven studies recognised racism as operating as an internalised experience. Overall, there were 26 studies defining racism at least two levels (structural/institutional/interpersonal/ internalised), with only two studies defining racism at all four levels. Three studies defined both direct and vicarious racial discrimination. More than half (n=270, 58%) of included studies referred to conceptual models, theories or frameworks related to racism and its influences on health outcomes (see Supplementary File 3).

Regarding the exposure measurement, almost all the studies (n=430, 93%) focused on child or youth self-reported experiences of direct, interpersonal racism. Experiences of direct racial discrimination were also reported by parents or teachers (n=41, 9%). Fewer studies focused on vicarious racism (n=21, 5%) or systemic, structural or institutional racism (n=28, 6%). The most common outcome was negative mental health (n=309, 67%), followed by health behaviours and sleep (n=126, 27%).

### Step two: Meta-analysis of physical health or biomarker outcomes (n=42)

The majority (n=41) of studies included in the meta-analysis measured the frequency of experiences of interpersonal racial discrimination and almost half (n=20) measured the contexts in which this occurred (e.g., at school, getting a job, getting medical care) (see Supplementary File 3). Most studies (n=40) used previously validated instruments to measure racial discrimination, which included the Experiences of Discrimination (EOD) scale (Krieger et al., 2005), the Everyday Discrimination Scale (EDS) (Williams et al., 1997), the Schedule of Racist Events (SRE) (Landrine & Klonoff, 1996), the Adolescent Discrimination Distress Index (ADDI) (Fisher et al., 2000), and the Daily Life Experience (DLE) scale (Seaton et al., 2009). The number of items used to measure racial discrimination ranged from 1 to 20. Response options were either binary (yes/no) or used the Likert scale (e.g., 3-point, 4-point, 5-point). When the exposure was measured in the association analyses, most studies (n=25) used continuous scores. The majority of studies measured racial discrimination with a time frame of either lifetime (n=24) or in the past year (n=13). More than half (n=25) of studies reported internal consistency of exposure measurement in the study sample, with Cronbach’s alpha ranging from 0.63 to 0.96.

Overall, the 42 studies reported a total of 83 associations with a range of physical health and biomarker indicators that were related to multiple physiological systems and processes. Outcomes were related to cardiometabolic risk (e.g. weight status, body composition, blood pressure, heart rate, pulse wave velocity, cardiac function), inflammation, endocrine function (e.g. pubertal timing, cortisol, dehydroepiandrosterone (DHEA)), respiratory function (e.g. asthma status, bronchodilator response), somatic symptoms, childhood illness and peak blood alcohol content.

Inflammation was the most commonly assessed group of outcomes (n=18) across a range of markers including CRP (n=5) and cytokines such as Interleukin (IL)-6 (n=3), IL-8 (n=3), IL-1β (n=3), Tumour Necrosis Factor (TNF)-α (n=3), and a composite measure of six cytokines (n=1). Weight status was next most commonly reported (n=13) followed by blood pressure (n=10), cortisol (n=8), asthma (n=6) and general somatic symptoms (n=6). Full details of outcome indicators are summarised in Supplementary File 4.

#### Inflammation

##### CRP

Five associations were reported between racism and CRP, with three examining maternal racial discrimination and two measuring child self-report racial discrimination. The majority of these were cross sectional (n=4). Most studies were conducted in the USA (n=4) and one in Australia. Latinx/Hispanic participants comprised most (70%) of the CRP study population, followed by Black participants (23%). Age at outcome assessment ranged from 4 to 20 years, with a focus on primary school and secondary school age. Three studies measured CRP in saliva and two in plasma/serum.

Overall, the pooled correlation coefficient between reported racism and CRP indicated a minimal to moderate positive association (*r*=0.07 [95% confidence interval (CI): 0.02, 0.12, p<0.01 (p value for the pooled effect size), k=5 (number of studies included in the meta-analysis); Q test=3.39, df=4, p=0.50 (p value for the homogeneity test)] (Figure 2A). However, when the pooled effect size was conducted within studies measuring CRP in saliva only (Figure 2B), we observed a marginal negative association, although the results were imprecise (*r*=-0.02 [95% CI: −0.15, 0.11, p=0.77; k=3; Q=0.93, df=2, p=0.63]).

**Figure 2.**
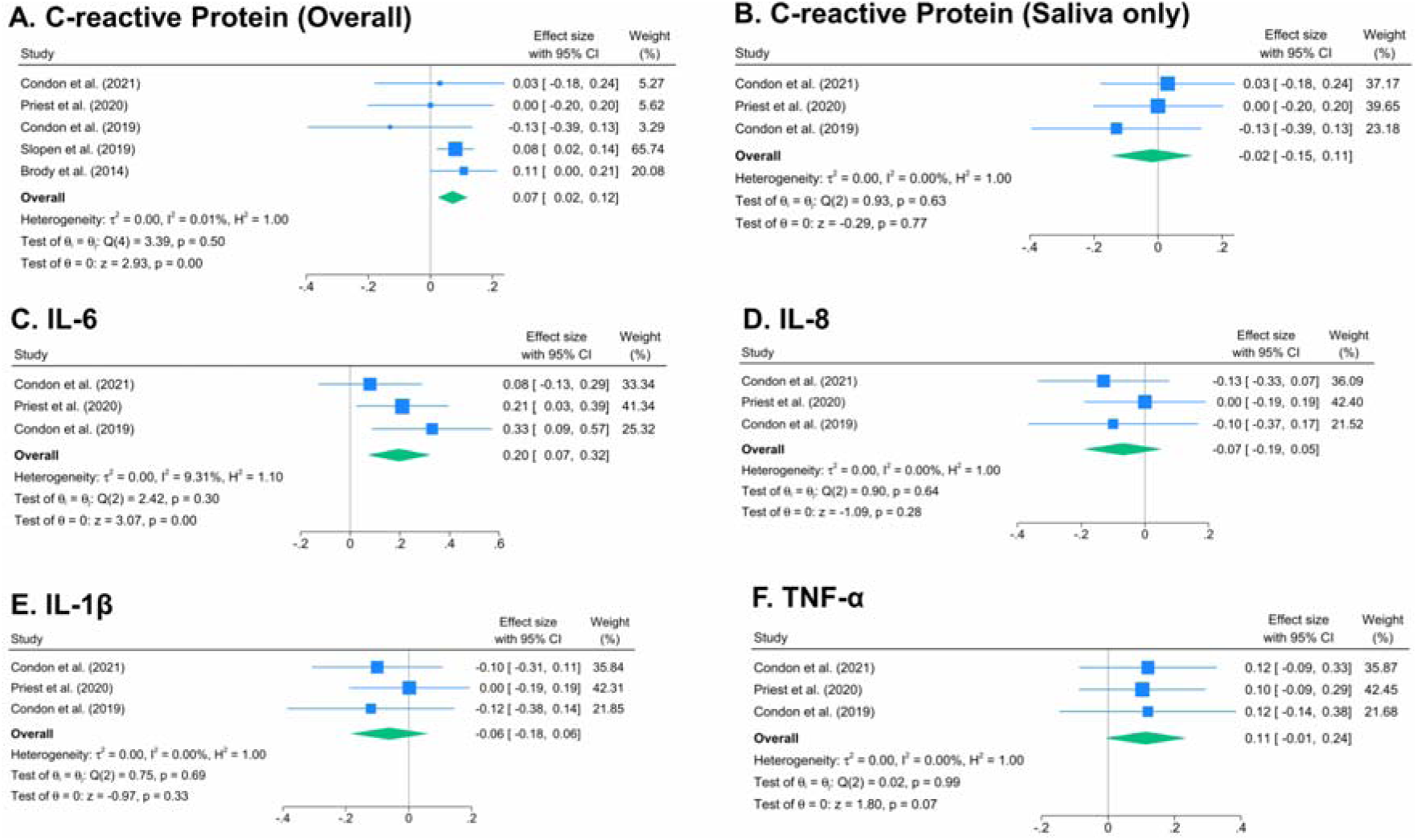
Associations between racism and A) C-reactive Protein (overall); B) C-reactive Protein (saliva only); C) IL-6; D) IL-8; E) IL-1β; and F) TNF-α.

#### Cytokines (IL-6, IL-8, IL-1***β***, and TNF-***α***)

There were three studies examining associations between racism and each of IL-6, IL-8, IL-1β, and TNF-α. Two studies examined maternal racial discrimination and one measured child self-report racial discrimination. All studies were cross sectional. Two studies were conducted in the USA and one study in Australia. Latinx/Hispanic participants comprised nearly one third (32%) of the pooled study population, followed by Black participants (22%) and white participants (20%). The remaining 26% of the pooled study population comprised Aboriginal and Torres Strait Islander people, from ethnic minority backgrounds, and individuals categorised as “other” or “mixed” racial groups. Age at outcome assessment ranged from 4 to 15 years, with a focus on primary school age. All studies measured cytokines in saliva.

The pooled correlation coefficient between reported racism and IL-6 was *r*=0.20 [95% CI: 0.07, 0.32, p<0.01; k=3; Q=2.42, df=2, p=0.30], suggesting that exposure to racism was moderately associated with higher levels of IL-6 (Figure 2C). The pooled correlation coefficient between reported racism and IL-8 and IL-1β indicated minimal negative associations: IL-8 (*r*=-0.07 [95% CI: −0.19, 0.05, p=0.28; k=3; Q=0.90, df=2, p=0.64]) (Figure 2D); IL-1β (*r*=-0.06 [95% CI: −0.18, 0.06, p=0.33; k=3; Q=0.75, df=2, p=0.69]) (Figure 2E), while exposure to racism appeared to be moderately associated with higher levels of TNF-α (*r*=0.11 [95% CI: −0.01, 0.24, p=0.07; k=3; Q=0.02, df=2, p=0.99] (Figure 2F). However, the results were imprecise with 95%CIs indicating both positive and negative associations for IL-8, IL-1β and TNF-α.

#### Weight status

##### BMI z-score

Five associations were reported between racism and BMI z-score. Three studies examined maternal racial discrimination and two measured child self-report racial discrimination. Age three studies were cross sectional and two were cohort. Most studies were conducted in the USA (n=4) and one study in Australia. White participants comprised nearly half (47%) of the pooled study population, followed by Latinx/Hispanic participants (30%) and Black participants (12%). Age at outcome assessment ranged from 3 to 16 years, with a focus on primary school age. All studies measured height and weight using objective scales. Overall, the pooled correlation coefficient between reported racism and BMI z-score was *r*=0.03 [95% CI: −0.05, 0.10, p=0.47; k=5; Q=10.30, df=4, p=0.04], suggesting a marginal positive association between racism and BMI z-score (Figure 3A).

**Figure 3.**
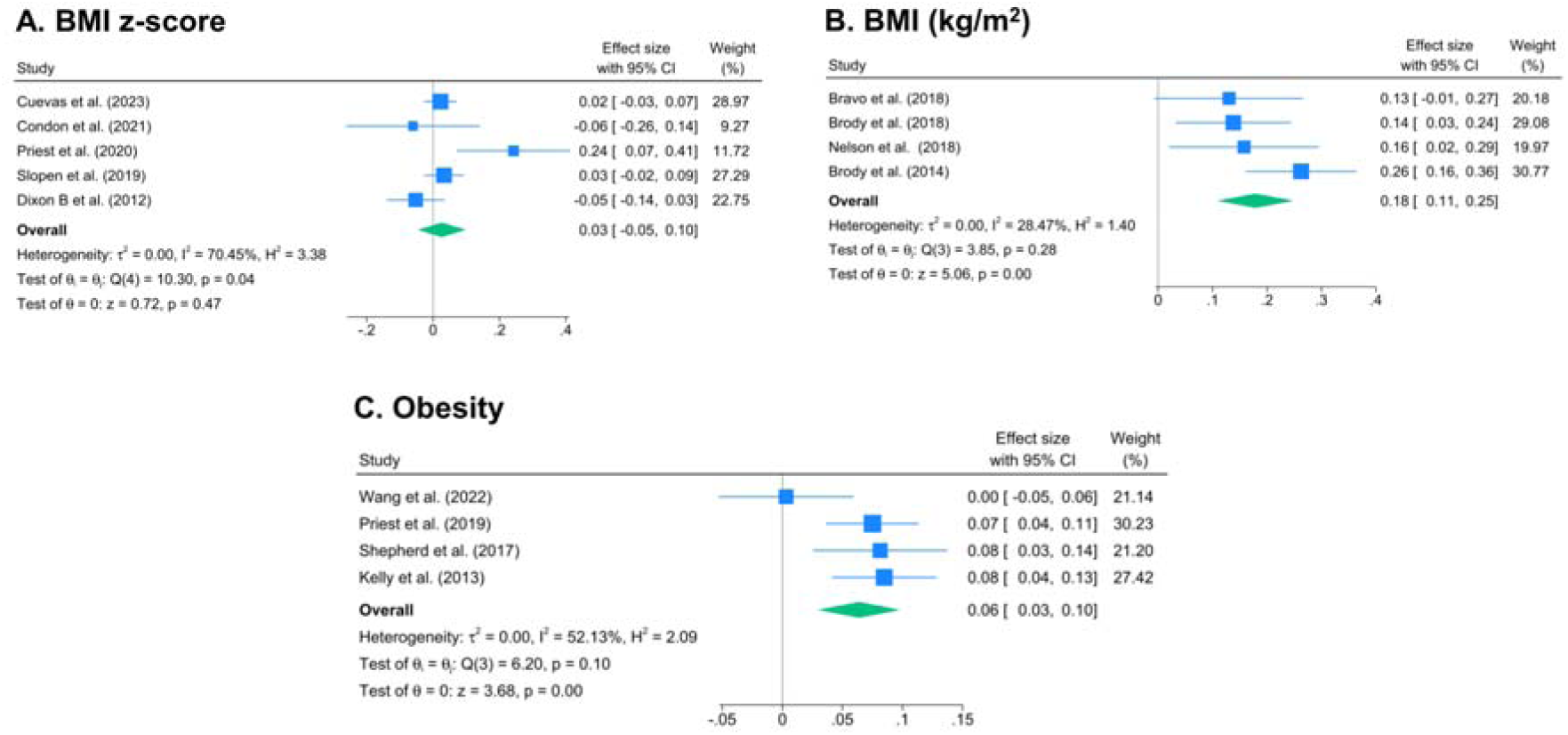
Associations between racism and A) Body mass index (BMI) z-score; B) BMI; and C) Obesity.

#### BMI (kg/m^2^)

Four associations were reported between racism and BMI (kg/m^2^) and all examined child self-report racial discrimination. Three studies were cohort and one cross sectional. All studies were conducted in the USA. Black participants comprised four fifths (80.9%) of the pooled study population, and the remaining participants were Latinx/Hispanic (19.1%). Age at outcome assessment ranged from 11 to 23 years, with a focus on post school age. Three studies measured height and weight using objective scales and one study was self-reported. Overall, the pooled correlation coefficient between reported racism and BMI (kg/m^2^) was *r*=0.18 [95% CI: 0.11, 0.25, p<0.01; k=4; Q=3.85, df=3, p=0.28], suggesting that exposure to racism was moderately associated with higher levels of BMI (kg/m^2^) (Figure 3B).

#### Obesity

Four associations were reported between racism and obesity status. Two studies examined parent-report racial discrimination, one assessed child self-report racial discrimination and one examined structural racism using the Black-White dissimilarity index. Three studies were cohort and one cross sectional. Two studies were conducted in Australia, one in the USA and one in the UK. Black participants comprised four fifths (46.9%) of the pooled study population, followed by white participants (29.5%) and Latinx/Hispanic participants (15.4%). Age at outcome assessment ranged from 5 to 17 years, with a focus on primary school age. Two studies measured obesity status using objective scales and two studies were self-reported. Overall, the pooled correlation coefficient between reported racism and obesity was *r*=0.06 [95% CI: 0.03, 0.10, p<0.01; k=4; Q=6.20, df=3, p=0.10], suggesting minimal to moderate positive associations (Figure 3C).

#### Blood pressure

There were five studies examining associations of racism with systolic blood pressure (SBP) and diastolic blood pressure (DBP) respectively. Three studies examined child self-report racial discrimination, one examined maternal racial discrimination and one indirect/ witnessing racial discrimination. Three studies were cross sectional, one was cohort and one experimental. Most studies (n=4) were conducted in the USA and one in Australia. Black participants comprised two thirds (67.7%) of the pooled study population, followed by Latinx/Hispanic participants (10.6%) and white participants (9.5%). Age at outcome assessment ranged from 4 to 21 years, with a focus on primary school and secondary school age. All studies measured SBP and DBP using objective blood pressure monitors.

Overall, the pooled correlation coefficient between reported racism and SBP was *r*=0.13 [95% CI: 0.05, 0.21, p<0.01; k=5; Q=5.64, df=4, p=0.23] (Figure 4A), suggesting minimal to moderate positive associations, while for DBP (Figure 4B) *r*=0.08 [95% CI: 0, 0.16, p=0.06; k=5; Q=6.41, df=4, p=0.17], suggesting a marginal positive association that was slightly imprecise.

**Figure 4.**
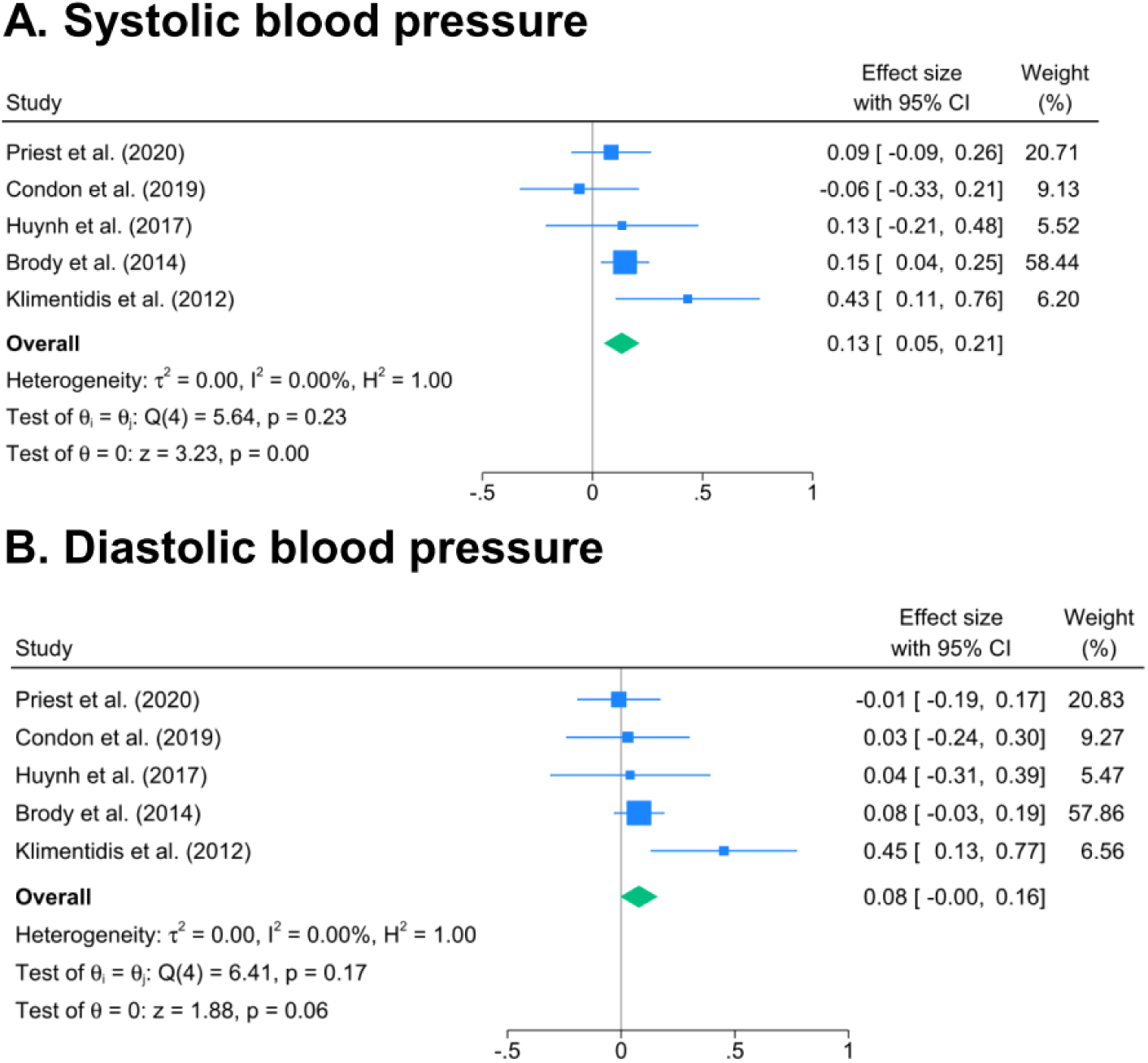
Associations between racism and A) Systolic blood pressure; and B) Diastolic blood pressure.

#### Cortisol

Eight associations were reported between racism and cortisol. Most (n=6) examined child self-report racial discrimination, one study measured maternal racial discrimination and one examined indirect/witnessing racial discrimination. Four studies were cross sectional, three were cohort and one experimental. All studies were conducted in the USA. Black participants comprised nearly three quarters (75.9%) of the cortisol study population, followed by Latinx/ Hispanic participants (19.8%) and white participants (4.3%). Age at outcome assessment ranged from 4 to 23 years, with a focus on post school age. Six studies measured cortisol in saliva, one study in urine and one in hair. Even across studies collecting salivary cortisol (n=6), assessment of cortisol varied, with four studies assessing cortisol at three or four time points during the day/interview, one measuring cortisol five times per day for three days, and one measuring cortisol at a single time point.

Overall, the pooled correlation coefficient between reported racism and cortisol was small: *r*=0.08 [95% CI: −0.02, 0.17, p=0.12; k=8; Q=17.30, df=7, p=0.02], with the 95% CI that was compatible with moderate positive associations or small negative associations (Figure 5A). However, when the pooled effect size was conducted within studies measuring cortisol in saliva only (Figure 5B), we estimated small to moderate positive associations, indicating that exposure to racism was associated with higher levels of cortisol (*r*=0.13 [95% CI: 0.02, 0.23, p=0.02; k=6; Q=10.67, df=5, p=0.06]).

**Figure 5.**
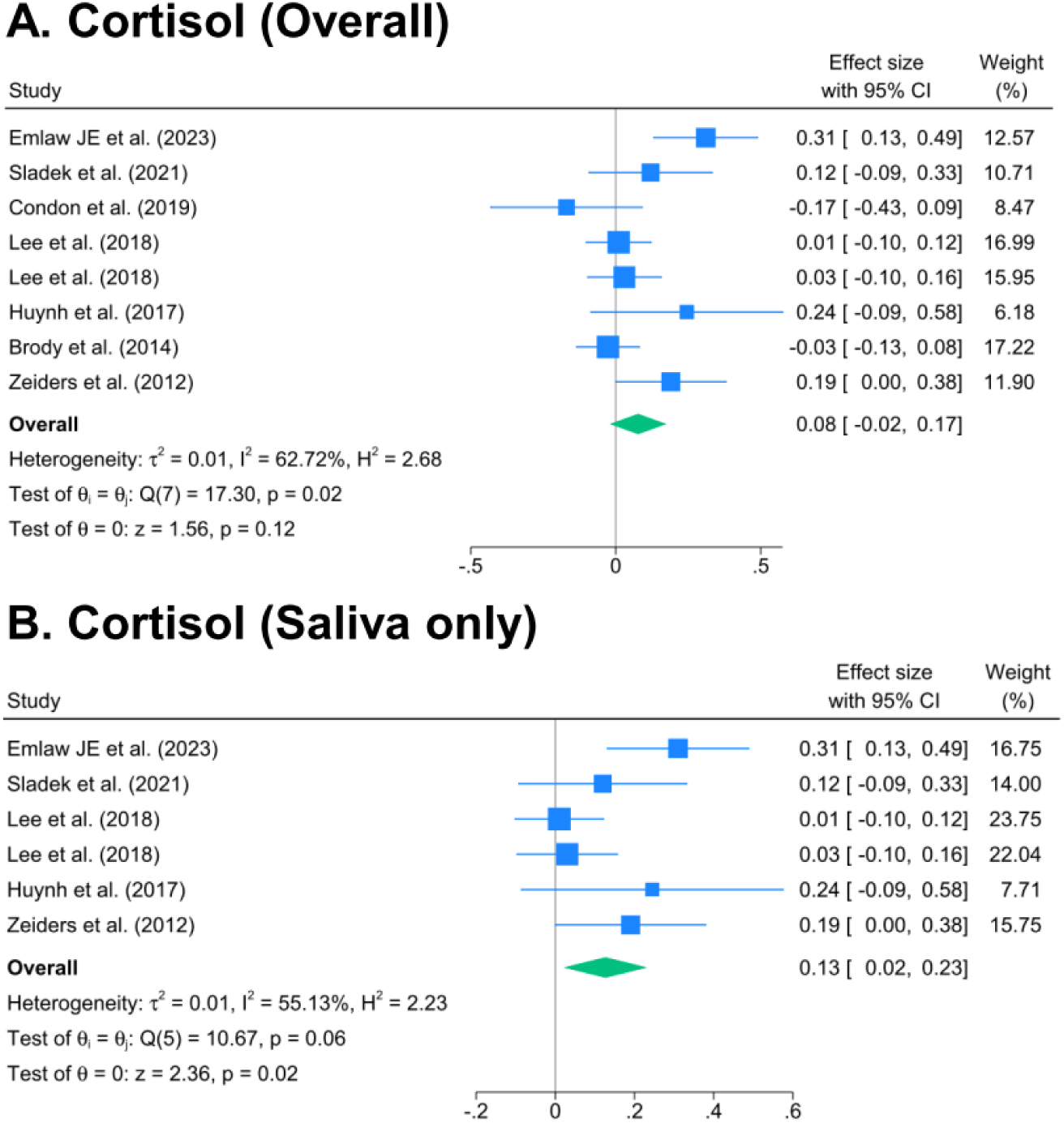
Associations between racism and A) Cortisol (overall); and B) Cortisol (saliva only).

#### Asthma

Six associations were reported between racism and asthma status. Three studies examined parent-report racial discrimination, two studies assessed child self-report racial discrimination and one examined structural racism using the Black-White dissimilarity index. Three studies were cohort, two were cross sectional, and one case-control. Two studies were conducted in the USA, one in the UK, one in Australia, one in New Zealand, and one in Brazil. Black participants comprised more than two fifths (46%) of the cortisol study population, followed by white participants (30%) and Latinx/Hispanic participants (16%). Aboriginal and Torres Strait Islander people, individuals from ethnic minority backgrounds, and individuals categorised as “other” or “mixed” racial groups together comprised the remaining 8.6% of the pooled study population. Age at outcome assessment ranged from 5 to 21 years, with a focus on primary school age. Asthma was defined by self-reported physician diagnosis in three studies, self-reported in two studies and one parent-report. Overall, the pooled correlation coefficient and the 95% CI indicated small positive associations between reported racism and asthma *r*=0.08 [95% CI: 0.03, 0.13, p<0.01; k=6; Q=45.63, df=5, p<0.01] (Figure 6).

**Figure 6.**
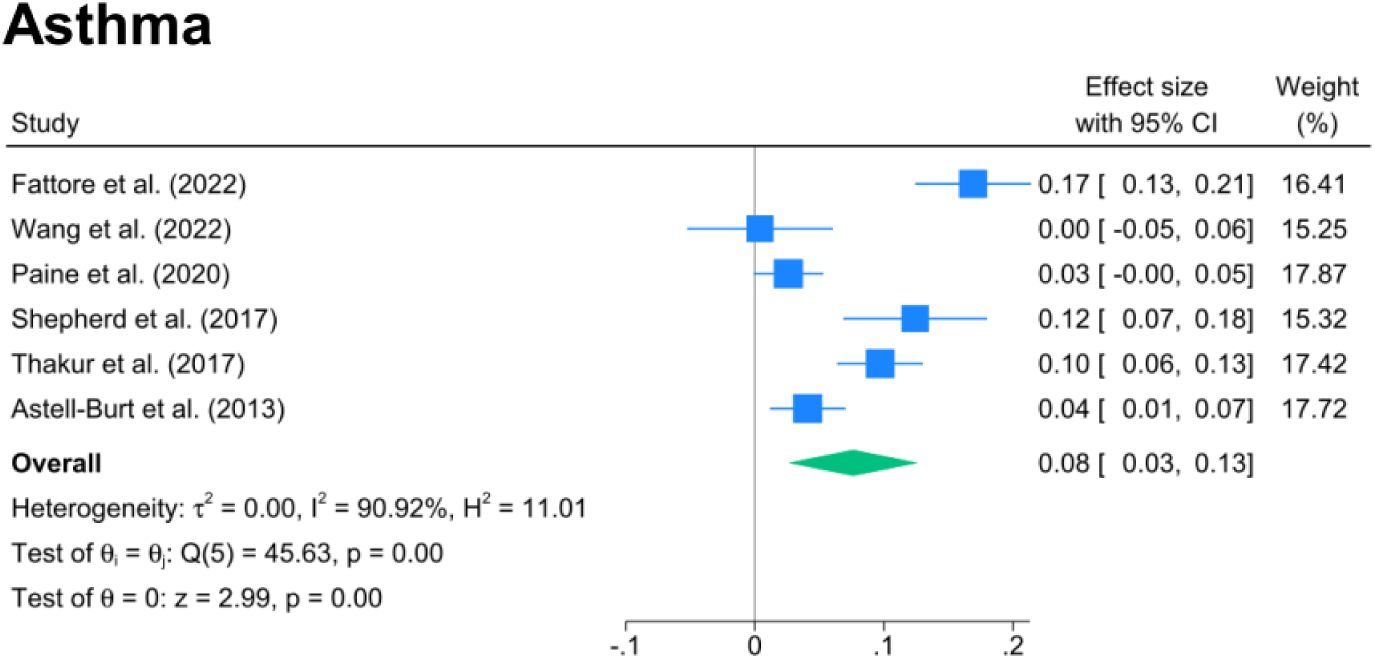
Associations between racism and asthma.

#### Somatic Symptoms

Six associations were reported between racism and somatic symptoms. All studies examined child self-report racial discrimination. Four studies were cohort and two were cross sectional. All studies were conducted in the USA. One study did not report the racial/ethnic composition. Among the remaining studies, Latinx/Hispanic participants comprised more than half (52.5%) of the pooled study population, followed by Asian participants (25.5%), Black participants (9.9%) and the remaining participants including white, “other” and “mixed” racial groups (12.1%). Age at outcome assessment ranged from 13 to 21 years, with a focus on secondary school age. Somatic symptoms were self-reported in all six studies. Overall, the pooled correlation coefficient between reported racism and cortisol indicated a moderate positive association *r*=0.20 [95% CI: 0.12, 0.28, p<0.01; k=6; Q=26.21, df=5, p<0.01], with the 95%CI also indicating small to moderate positive associations (Figure 7).

**Figure 7.**
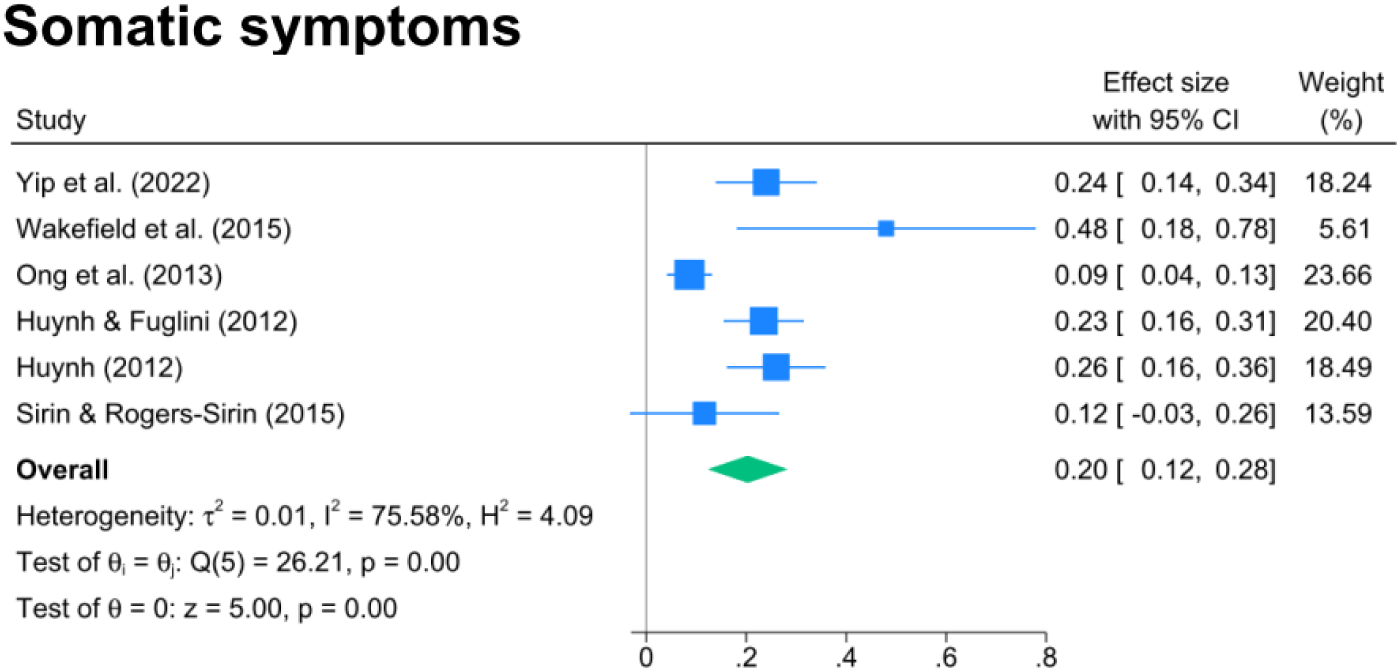
Associations between racism and somatic symptoms.

#### Sensitivity analyses results

When using maximally adjusted estimates to calculate the overall effect size, we found similar results across outcome indicators (see Supplementary File 5).

#### Mediators and moderators between racism and physical health or biological markers

Of the 42 studies examining physical health and biomarker outcomes, only three examined potential mediators (e.g., depressive symptoms, anxiety, social stress) linking racial discrimination with a specific outcome indicator and 18 studies examined potential moderators (e.g., race, neighbourhood racial composition, racial identity) of the relationship between racial discrimination and a specific outcome indicator (see Supplementary File 3 for full details). However, due to the high heterogeneity in the measures used for mediators and moderators, a pooled effect size was not generated.

#### Quality assessment and publication bias

Among the 42 studies on physical health and biological markers, the most common study design was longitudinal (n=20), followed by cross-sectional (n=19). Two studies used case-control designs and only one study was experimental (see Supplementary File 2 for details). The majority of longitudinal studies had fair (65%, 13/20) or good (10%, 2/20) quality. As for cross-sectional studies, most studies had good (63.2%, 12/19) or fair (26.3%, 5/19) quality. Regarding the two case-control studies and the one experimental study, we found all had good quality.

The funnel plots (see Supplementary File 6) and results from Egger’s tests to assess funnel plot asymmetry suggest little to weak evidence of publication bias: cortisol (z=0.84, p=0.40), CRP (z=-1.40, p=0.16), IL-1β (z=-0.66, p=0.51), IL-6 (z=0.59, p=0.55), IL-8 (z=-0.47, p=0.64), TNF-α (z=0.09, p=0.93), asthma (z=0.51, p=0.61), somatic symptoms (z=1.88, p=0.06), SBP (z=0.38, p=0.71), DBP (z=0.78, p=0.44), BMI z-score (z=0.56, p=0.57), BMI (z=-0.94, p=0.34), and obesity (z=-0.97, p=0.33).

## DISCUSSION

Our review organises and synthesises recent evidence on the extent to which racism is associated with health and wellbeing among children and youth, updating the previous systematic review (Priest et al., 2013). We hope this informs future research priorities and motivates strategic action to address systemic racism and its harms for children and youth. The field of racism and child and youth health has substantially grown since the original review, with an almost fourfold increase in the number of studies published. Yet there remains an ongoing need within studies for far more explicit engagement with theoretical frameworks of racism and health, including defining and conceptualising racism and the ways in which racism as a system of oppression impacts child and youth health. While empirical attention to the impacts of systemic, structural and institutional racism on child and youth health has increased in recent years, it still remains relatively small. Overwhelmingly, the field continues to focus on direct interpersonal experiences of racism. Attention to health outcomes beyond mental health and health behaviours, to elucidating the pathways by which racism becomes embodied and biologically expressed during childhood and youth, and to racism’s impacts in early and middle childhood also remain outstanding priorities. Physical health and biomarker outcomes have received more attention in recent years though are still less than ten per cent of all studies identified. Our meta-analysis found minimal to moderate positive associations between racism and increased inflammation as measured by CRP and IL-6, higher levels of BMI and obesity, higher levels of systolic blood pressure and salivary cortisol, asthma and somatic symptoms. There were marginal positive associations between racism and increased inflammation measured by TNF-α, higher levels of cortisol, BMI-z score and diastolic blood pressure, with imprecise estimates and wide CIs. We found minimal negative associations of racism with IL-1β and IL-8, with imprecise estimates and wide CIs. Together these findings suggest that racism is associated with multiple physiological systems and biological processes in childhood and adolescence. This has implications for health and wellbeing during childhood and adolescence, as well as for future chronic disease risk.

Consistent with previous systematic reviews (Benner et al., 2018; Priest et al., 2013), we found that most included studies were conducted in the USA, particularly in urban contexts, suggesting a need for more evidence from other countries and contexts. Critically, only 14% of studies included Indigenous populations, reinforcing calls for far greater attention to the health impacts of racism for Indigenous children and youth as distinct from other racialised groups and as deeply embedded within settler-colonialism (Uink et al., 2022). Although more studies have larger sample sizes (n ≥1000) which can improve statistical power and enable more precise estimates (Fraley & Vazire, 2014), there remains a predominance of convenience sampling, which creates generalizability issues for statistical studies. In this review, more than two-fifths of included studies were longitudinal, double that of the previous review (Priest et al., 2013). This reflects a growing recognition of the need to understand the dynamics of variables over time and the long-term effect of racism on health and wellbeing over the life course.

Racism and racial discrimination were formally defined in fewer than two fifths of the studies examined. The explicit use of the term ‘racism’ and the explicit conceptualisations and definitions of racism as systemic and structural, including as the driver of racial discrimination enacted between individuals, is an identified priority for studies of racism and health despite firm opposition in some scientific contexts (Howe et al., 2022; Krieger et al., 2021; Michaels et al., 2023; Wray-Lake et al., 2022; Zambrana & Williams, 2022). Only just over half of the included studies presented the theoretical models, theories, or frameworks underpinning their research. Yet the opportunities to produce more meaningful evidence offered by the improved methodological rigour of the last decade can only go so far without its location in appropriate theory, including those related to racism and health, racialised health inequities and embodiment, and socio-ecological models of child and youth health and development (Javadi et al., 2023; Krieger, 2012; Slopen & Heard-Garris, 2022; Williams & Mohammed, 2013).

Our findings that racism is associated with multiple negative physical health and biomarker outcomes are consistent with previous reviews (Cuevas et al., 2020; Lawrence et al., 2022). One narrative review identified associations between discrimination and inflammatory markers including CRP and IL-6, though did not report findings by type of discrimination or inflammatory marker used (Cuevas et al., 2020). A systematic review and meta-analysis of the Everyday Discrimination Scale and biomarker outcomes also found minimal to moderate positive associations between discrimination and elevated CRP though unlike this present review did not find minimal to moderate positive associations between discrimination and IL-6 (Lawrence et al., 2022). This may be related to different exposure measures, age of participants, measurement approaches including plasma or saliva, or other study characteristics. Unlike our meta-analysis findings that found moderate evidence of an association between racism and salivary cortisol, previous reviews have reported a lack of evidence for association between the Everyday Discrimination Scale and cortisol (Lawrence et al., 2022) and between racial discrimination and cortisol (Korous et al., 2017). Consistently across all reviews, including this present one, is the considerable variation in measurement approaches of cortisol and the complexity in measuring discrimination and cortisol. Our findings in relation to systolic blood pressure, weight status, asthma and somatic symptoms are also consistent with evidence among adults that shows the harmful impacts of racism on these outcomes (Paradies et al., 2015) and with evidence regarding the impacts of childhood adversity more generally on these outcomes in childhood and adolescence (Oh et al., 2018; Suglia et al., 2020).

While there is variation in the effect sizes and precision across outcome indicators, together the findings across outcomes suggest racism exposure is associated with multiple biological disruptions for children and youth and that these relate to a wide range of physiological processes including immune, endocrine, respiratory and cardiovascular function. This is consistent with conceptual and empirical evidence regarding the highly integrated responses to harmful social and environmental exposures that occur across nervous, endocrine, and immune systems and the short and long term impacts of chronic activation of these systems in childhood (Danese & McEwen, 2012; Shonkoff et al., 2012). For example, increased overweight and obesity risk in response to racism may be related to structural and functional nervous system changes that can lead to behavioural responses such as physical activity levels, eating behaviours, as well as endocrine changes linked to chronic activation of the hypothalamic-pituitary-adrenal (HPA) axis such as increased cortisol and glucocorticoid resistance. Blood pressure increases can be related to dysregulation of the autonomic nervous system. Increased inflammation can also be related to changes in the autonomic nervous system, as well as to weight status, physical activity and dietary quality (Danese & McEwen, 2012; Shonkoff et al., 2012). It is well established that childhood and adolescence are life periods during which these biological systems are especially sensitive to social and environmental influences and that resulting structural changes and physiological disruptions can have lifelong implications (Patton et al., 2016; Shonkoff et al., 2021). Systemic racism fundamentally shapes these social and environmental influences acting as a distinct and substantial threat to the foundations of optimal health and development for children and youth from racialised communities (Shonkoff et al., 2021; Slopen & Heard-Garris, 2022).

The variability in the effects of racism across different outcome indicators identified in this review might be attributed to several factors. Methodological differences across studies, including sample size, racism exposure measures, approaches of physical health and biomarker measure and analysis, and analytic approaches including confounder selection and unmeasured covariates, may also account for inconsistencies in the findings. Together with key issues related to racism exposure measurement such as capturing timing, intensity, chronicity, and form of exposure, as well as potential mediating and moderating factors, age at outcome assessment may also explain variability in findings. For example, effects of childhood adversity on inflammation have been shown to differ by age of outcome, with stronger evidence of associations in late childhood compared to mid childhood possibly due to latency periods between exposure and outcome or that current inflammatory biomarkers are not sensitive to chronic inflammation in early life (O’Connor et al., 2020). With only three studies included in the meta-analysis examining moderating or mediating factors between racism and physical health and biomarker outcomes, this is an important area of future work.

Outcome measure selection and analytic approach which are also highlighted in previous reviews likely played a role in the variation across findings, particularly for inflammation and cortisol (Cuevas et al., 2020; Korous et al., 2017; Lawrence et al., 2022). Measurement of inflammatory markers in this review spanned saliva and plasma/serum though these may not be comparable with inflammation in saliva and are possibly a poor indicator of systemic (rather than intra-oral) inflammation (Priest et al., 2020). Further, CRP is an acute phase reactant and is considered a less than optimal marker of childhood chronic inflammation (Collier et al., 2019). Cytokines such as IL-6, IL-1β, and IL-8 also can each be associated with acute and chronic inflammatory responses, both alloimmune (non-self) and auto-immune threats and are associated with different disease states, e.g. IL-6, IL-8 with obesity, TNF-α with depression (Chen et al., 2017). A more promising inflammatory marker for capturing cumulative, chronic inflammation in childhood may be glycoprotein acetyls (GlycA), a nuclear magnetic resonance derived metabolite (Collier et al., 2019) with evidence of associations in childhood with adverse psychosocial exposures (O’Connor et al., 2020). There is also a need to conceptualise and quantify inflammation not as a static phenomenon captured by a single biomarker at one time point, but as a dynamic interaction of pro-inflammatory and anti-inflammatory pathways, and in the context of a persistent immune phenotype (Gamage et al., under review; McDade, 2023). The assessment of cortisol varied across saliva, hair and urine samples and included different timing of collection ranging from three or four time points during the day/interview, five times per day for three days, or a single time point. Each of these likely captures different aspects of the physiological stress response (Korous et al., 2017), making meaningful interpretation of these findings difficult.

Overall, our review showed that racism is associated with multiple poor physiological systems and biological processes in childhood and adolescence, although evidence remains relatively emergent. Nonetheless, these findings have implications for health and wellbeing during childhood and adolescence, as well as for future chronic disease risk, and begin to elucidate how racism becomes embodied and embedded in early life. Grounded in strong explicit conceptual and theoretical understandings of systemic racism and health combined with socio-ecological models of child and youth development, we must continue to highlight the profound health harms of racism for children and youth from racialised backgrounds. This includes countering false narratives implicitly or explicitly focused on individual behaviours or limitations of parents or children and young people and challenging narrow approaches that focus on childhood adversity and either add racial discrimination as simply another form of adversity within cumulative risk scores or ignore it altogether, instead of recognising racism as a fundamental and unique social force that shapes and drives other forms of childhood adversity. We must find new ways to bring together expertise across disciplines to ensure robust and comprehensive measurement of systemic racism exposure that is age, population and context appropriate, and to harness the best of biomarker and clinical research to ensure outcome measurement and interpretation is biologically and clinically meaningful and ultimately contributes to informing interventions to counter racism and its harmful health effects. Critically, this must be governed by and in authentic partnership with community organisations, children and young people and their families, and researchers from racialised communities (Australian Institute of Aboriginal and Torres Strait Islander Studies, 2020; Wray-Lake et al., 2022).

### Limitations

There were several limitations that should be noted. We only included eligible studies that were published in English from four online databases, which might miss studies published in other languages or the grey literature. Measurement bias with respect to specific outcome indicators and exposure may still exist in the studies used for the meta-analysis. Due to the small number of studies examining each outcome indicator, different exposure measures were included in each estimation of the pooled correlation coefficients, which may have introduced bias as associations may well vary by exposure measure. In addition, in the process of effect size conversion, formulas used to transform binary effect measures to correlation coefficients were based on approximations. Given that correlation coefficients were used to measure the effect size, results are likely to be subjected to confounding, however the sensitivity analysis with adjusted results indicated that the degree of confounding might be small. The majority of included studies in the meta-analysis used convenience samples which may have introduced selection bias. They were also predominantly conducted in the USA which may limit the generalisability of our findings to other geographical contexts and populations though it is unlikely with the harmful effects of childhood adversity well established across contexts (Soares et al., 2021).

## CONCLUSION

We found that racism was associated with a range of negative physical health and biomarker outcomes that relate to multiple physiological systems and biological processes in childhood and adolescence. These findings have implications for health and wellbeing during childhood and adolescence, as well as for future chronic disease risk, and begin to elucidate how systemic racism becomes embodied and embedded in early life. While more work is needed to understand these pathways and mechanisms, this must not be at the expense of, or considered necessary for action to address racism as a fundamental cause of child and youth health. We must move beyond description and enact collective and structural changes through policies, education, advocacy, and partnerships to eliminate racism and create a healthy and equitable future for all children and youth.

## Supporting information

PRISMA_2020_checklist

MOOSE_Checklist

## Data Availability

All data used in this manuscript are available in Supplementary files.

## Acknowledgment

This work was supported by the Victorian Government’s Operational Infrastructure Support Program.

## Author Contributions

Naomi Priest: Conceptualization, Methodology, Investigation, Supervision, Writing – Original Draft, Writing - Review & Editing; Chiao Kee Lim: Data curation, Formal analysis, Investigation, Validation, Writing - Review & Editing; Kate Doery: Conceptualization, Methodology, Investigation, Writing - Review & Editing; Jourdyn A. Lawrence: Conceptualization, Methodology, Investigation, Validation, Writing - Review & Editing; Georgia Zoumboulis: Investigation, Writing - Review & Editing; Gabriella King: Investigation, Writing - Review & Editing; Dewan Lamisa: Investigation, Writing - Review & Editing; Fan He: Investigation, Writing - Review & Editing; Rushani Wijesuriya: Methodology, Writing - Review & Editing; Camila M. Mateo: Investigation, Writing - Review & Editing; Shiau Chong: Investigation, Writing - Review & Editing; Mandy Truong: Investigation, Writing - Review & Editing; Ryan Perry: Investigation, Validation, Writing - Review & Editing; Paula Toko King: Investigation, Writing - Review & Editing; Natalie Paki Paki: Investigation, Writing - Review & Editing; Corey Joseph: Investigation, Writing - Review & Editing; Dot Pagram: Investigation, Writing - Review & Editing; Roshini Balasooriya Lekamge: Investigation, Writing - Review & Editing; Gosia Mikolajczak: Investigation, Writing - Review & Editing; Emily Darnett: Investigation, Writing - Review & Editing; Brigid Trenerry: Investigation, Writing - Review & Editing; Shloka Jha: Investigation, Writing - Review & Editing; Joan Gakii Masunga: Investigation, Writing - Review & Editing; Yin Paradies: Investigation, Writing - Review & Editing; Investigation, Writing - Review & Editing; Yvonne Kelly: Conceptualization, Writing - Review & Editing; Saffron Karlsen: Conceptualization, Writing - Review & Editing; Shuaijun Guo: Conceptualization, Data curation, Methodology, Project administration, Formal analysis, Investigation, Writing – Original Draft, Writing - Review & Editing.

