## Supplementary material for "Racism and health and wellbeing among children and youth - an updated systematic review and meta-analysis": Supplementary file 1. Detailed search strategies.docx

**Supplementary File 1**. Detailed search strategies in each database

**PubMed**

PubMed encompasses peer-reviewed journal articles and online books, covering the fields of biomedicine and health (Williamson & Minter, 2019).

((("racism"[Title/Abstract] OR "racial-discriminat*"[Title/Abstract] OR "racial-prejudice"[Title/Abstract] OR "racist-event*"[Title/Abstract] OR "racist-episode*"[Title/Abstract] OR "racial-stereotype*"[Title/Abstract] OR "race-related-stress"[Title/Abstract]) OR ("unfair*"[Title/Abstract] AND "treat*"[Title/Abstract] AND ("race"[Title/Abstract] OR "racial*"[Title/Abstract] OR "ethnic*"[Title/Abstract] OR "cultur*"[Title/Abstract] OR "religio*"[Title/Abstract] OR "migrant*"[Title/Abstract] OR "refugee*"[Title/Abstract] OR "asylum"[Title/Abstract])) OR (("discriminat*"[Title/Abstract] OR "bias*"[Title/Abstract] OR "prejudic*"[Title/Abstract] OR "hostil*"[Title/Abstract] OR "harass*"[Title/Abstract] OR "bully*"[Title/Abstract] OR "cyberbull*"[Title/Abstract] OR "cyber-bull*"[Title/Abstract] OR "oppress*"[Title/Abstract]) AND ("race"[Title/Abstract] OR "racial*"[Title/Abstract] OR "ethnic*"[Title/Abstract] OR "cultur*"[Title/Abstract] OR "religio*"[Title/Abstract] OR "migrant*"[Title/Abstract] OR "refugee*"[Title/Abstract] OR "asylum"[Title/Abstract]))) AND ("newborn*"[Title/Abstract] OR "new-born*"[Title/Abstract] OR "baby"[Title/Abstract] OR "babies"[Title/Abstract] OR "neonat*"[Title/Abstract] OR "neo-nat*"[Title/Abstract] OR "infan*"[Title/Abstract] OR "toddler*"[Title/Abstract] OR "pre-schooler*"[Title/Abstract] OR "preschooler*"[Title/Abstract] OR "kinder"[Title/Abstract] OR "kinders"[Title/Abstract] OR "kindergarten*"[Title/Abstract] OR "boy"[Title/Abstract] OR "boys"[Title/Abstract] OR "girl"[Title/Abstract] OR "girls"[Title/Abstract] OR "child"[Title/Abstract] OR "children"[Title/Abstract] OR "childhood"[Title/Abstract] OR "pediatric*"[Title/Abstract] OR "paediatric*"[Title/Abstract] OR "adolescen*"[Title/Abstract] OR "youth"[Title/Abstract] OR "youths"[Title/Abstract] OR "teen"[Title/Abstract] OR "teens"[Title/Abstract] OR "teenage*"[Title/Abstract] OR "school-age*"[Title/Abstract] OR "schoolage*"[Title/Abstract] OR "school-child*"[Title/Abstract] OR "schoolchild*"[Title/Abstract] OR "school-girl*"[Title/Abstract] OR "schoolgirl*"[Title/Abstract] OR "school-boy*"[Title/Abstract] OR "schoolboy*"[Title/Abstract] OR "young-person*"[Title/Abstract] OR "young-people"[Title/Abstract]) AND ("health-care"[Title/Abstract] OR "healthcare"[Title/Abstract] OR "health-service*"[Title/Abstract] OR "clinic"[Title/Abstract] OR "clinics"[Title/Abstract] OR "ill-health"[Title/Abstract] OR "wellbeing"[Title/Abstract] OR "well-being"[Title/Abstract] OR "disease*"[Title/Abstract] OR "illness*"[Title/Abstract] OR "bmi"[Title/Abstract] OR "body-mass-index"[Title/Abstract] OR "anthropometric*"[Title/Abstract] OR "WHR"[Title/Abstract] OR "waist-hip-ratio"[Title/Abstract] OR "cardiovascular"[Title/Abstract] OR "cardio-vascular"[Title/Abstract] OR "hypertension"[Title/Abstract] OR "blood-pressure"[Title/Abstract] OR "cardiometabolic"[Title/Abstract] OR "cardio-metabolic"[Title/Abstract] OR "biomarker*"[Title/Abstract] OR "biological-marker*"[Title/Abstract] OR "obese"[Title/Abstract] OR "obesity"[Title/Abstract] OR "overweight"[Title/Abstract] OR "depress*"[Title/Abstract] OR "anxiety"[Title/Abstract] OR "anxious*"[Title/Abstract] OR "mental-health"[Title/Abstract] OR "mental-disorder*"[Title/Abstract] OR "stress"[Title/Abstract] OR "distress*"[Title/Abstract] OR "suicid*"[Title/Abstract] OR "sleep"[Title/Abstract] OR "psychosis"[Title/Abstract] OR "tobacco"[Title/Abstract] OR "smoke*"[Title/Abstract] OR "smoking"[Title/Abstract] OR "drug*"[Title/Abstract] OR "alcohol*"[Title/Abstract] OR "substance-use"[Title/Abstract] OR "substance-related-disorder*"[Title/Abstract] OR "resilien*"[Title/Abstract] OR "self-esteem"[Title/Abstract] OR "self-worth"[Title/Abstract] OR "self-concept"[Title/Abstract] OR "quality-of-life"[Title/Abstract] OR "life-satisfaction"[Title/Abstract] OR "personal-satisfaction"[Title/Abstract] OR "conduct-disorder*"[Title/Abstract] OR "aggression"[Title/Abstract] OR "aggressive*"[Title/Abstract] OR (("social"[Title/Abstract] OR "behavio*"[Title/Abstract] OR "emotion*"[Title/Abstract] OR "developmental*"[Title/Abstract] OR "psychological*"[Title/Abstract] OR "learning*"[Title/Abstract]) AND ("difficul*"[Title/Abstract] OR "problem*"[Title/Abstract] OR "delay*"[Title/Abstract] OR "adjust*"[Title/Abstract] OR "adapt*"[Title/Abstract])) OR (("pregnancy"[Title/Abstract] OR "birth"[Title/Abstract] OR "gestation*"[Title/Abstract]) AND ("outcome*"[Title/Abstract] OR "preterm"[Title/Abstract] OR "pre-term"[Title/Abstract] OR "premature"[Title/Abstract] OR "small-for-gestational-age"[Title/Abstract])) OR "low-birthweight"[Title/Abstract] OR "low-birth-weight"[Title/Abstract]) AND (NOTNLM OR publisher[sb] OR inprocess[sb] OR pubmednotmedline[sb] OR indatareview[sb] OR pubstatusaheadofprint)) NOT (((("racism"[Title/Abstract] OR "racial-discriminat*"[Title/Abstract] OR "racial-prejudice"[Title/Abstract] OR "racist-event*"[Title/Abstract] OR "racist-episode*"[Title/Abstract] OR "racial-stereotype*"[Title/Abstract] OR "race-related-stress"[Title/Abstract]) OR ("unfair*"[Title/Abstract] AND "treat*"[Title/Abstract] AND ("race"[Title/Abstract] OR "racial*"[Title/Abstract] OR "ethnic*"[Title/Abstract] OR "cultur*"[Title/Abstract] OR "religio*"[Title/Abstract] OR "migrant*"[Title/Abstract] OR "refugee*"[Title/Abstract] OR "asylum"[Title/Abstract])) OR (("discriminat*"[Title/Abstract] OR "bias*"[Title/Abstract] OR "prejudic*"[Title/Abstract] OR "hostil*"[Title/Abstract] OR "harass*"[Title/Abstract] OR "bully*"[Title/Abstract] OR "cyberbull*"[Title/Abstract] OR "cyber-bull*"[Title/Abstract] OR "oppress*"[Title/Abstract]) AND ("race"[Title/Abstract] OR "racial*"[Title/Abstract] OR "ethnic*"[Title/Abstract] OR "cultur*"[Title/Abstract] OR "religio*"[Title/Abstract] OR "migrant*"[Title/Abstract] OR "refugee*"[Title/Abstract] OR "asylum"[Title/Abstract]))) AND ("newborn*"[Title/Abstract] OR "new-born*"[Title/Abstract] OR "baby"[Title/Abstract] OR "babies"[Title/Abstract] OR "neonat*"[Title/Abstract] OR "neo-nat*"[Title/Abstract] OR "infan*"[Title/Abstract] OR "toddler*"[Title/Abstract] OR "pre-schooler*"[Title/Abstract] OR "preschooler*"[Title/Abstract] OR "kinder"[Title/Abstract] OR "kinders"[Title/Abstract] OR "kindergarten*"[Title/Abstract] OR "boy"[Title/Abstract] OR "boys"[Title/Abstract] OR "girl"[Title/Abstract] OR "girls"[Title/Abstract] OR "child"[Title/Abstract] OR "children"[Title/Abstract] OR "childhood"[Title/Abstract] OR "pediatric*"[Title/Abstract] OR "paediatric*"[Title/Abstract] OR "adolescen*"[Title/Abstract] OR "youth"[Title/Abstract] OR "youths"[Title/Abstract] OR "teen"[Title/Abstract] OR "teens"[Title/Abstract] OR "teenage*"[Title/Abstract] OR "school-age*"[Title/Abstract] OR "schoolage*"[Title/Abstract] OR "school-child*"[Title/Abstract] OR "schoolchild*"[Title/Abstract] OR "school-girl*"[Title/Abstract] OR "schoolgirl*"[Title/Abstract] OR "school-boy*"[Title/Abstract] OR "schoolboy*"[Title/Abstract] OR "young-person*"[Title/Abstract] OR "young-people"[Title/Abstract]) AND ("health-care"[Title/Abstract] OR "healthcare"[Title/Abstract] OR "health-service*"[Title/Abstract] OR "clinic"[Title/Abstract] OR "clinics"[Title/Abstract] OR "ill-health"[Title/Abstract] OR "wellbeing"[Title/Abstract] OR "well-being"[Title/Abstract] OR "disease*"[Title/Abstract] OR "illness*"[Title/Abstract] OR "bmi"[Title/Abstract] OR "body-mass-index"[Title/Abstract] OR "anthropometric*"[Title/Abstract] OR "WHR"[Title/Abstract] OR "waist-hip-ratio"[Title/Abstract] OR "cardiovascular"[Title/Abstract] OR "cardio-vascular"[Title/Abstract] OR "hypertension"[Title/Abstract] OR "blood-pressure"[Title/Abstract] OR "cardiometabolic"[Title/Abstract] OR "cardio-metabolic"[Title/Abstract] OR "biomarker*"[Title/Abstract] OR "biological-marker*"[Title/Abstract] OR "obese"[Title/Abstract] OR "obesity"[Title/Abstract] OR "overweight"[Title/Abstract] OR "depress*"[Title/Abstract] OR "anxiety"[Title/Abstract] OR "anxious*"[Title/Abstract] OR "mental-health"[Title/Abstract] OR "mental-disorder*"[Title/Abstract] OR "stress"[Title/Abstract] OR "distress*"[Title/Abstract] OR "suicid*"[Title/Abstract] OR "sleep"[Title/Abstract] OR "psychosis"[Title/Abstract] OR "tobacco"[Title/Abstract] OR "smoke*"[Title/Abstract] OR "smoking"[Title/Abstract] OR "drug*"[Title/Abstract] OR "alcohol*"[Title/Abstract] OR "substance-use"[Title/Abstract] OR "substance-related-disorder*"[Title/Abstract] OR "resilien*"[Title/Abstract] OR "self-esteem"[Title/Abstract] OR "self-worth"[Title/Abstract] OR "self-concept"[Title/Abstract] OR "quality-of-life"[Title/Abstract] OR "life-satisfaction"[Title/Abstract] OR "personal-satisfaction"[Title/Abstract] OR "conduct-disorder*"[Title/Abstract] OR "aggression"[Title/Abstract] OR "aggressive*"[Title/Abstract] OR (("social"[Title/Abstract] OR "behavio*"[Title/Abstract] OR "emotion*"[Title/Abstract] OR "developmental*"[Title/Abstract] OR "psychological*"[Title/Abstract] OR "learning*"[Title/Abstract]) AND ("difficul*"[Title/Abstract] OR "problem*"[Title/Abstract] OR "delay*"[Title/Abstract] OR "adjust*"[Title/Abstract] OR "adapt*"[Title/Abstract])) OR (("pregnancy"[Title/Abstract] OR "birth"[Title/Abstract] OR "gestation*"[Title/Abstract]) AND ("outcome*"[Title/Abstract] OR "preterm"[Title/Abstract] OR "pre-term"[Title/Abstract] OR "premature"[Title/Abstract] OR "small-for-gestational-age"[Title/Abstract])) OR "low-birthweight"[Title/Abstract] OR "low-birth-weight"[Title/Abstract]) AND (NOTNLM OR publisher[sb] OR inprocess[sb] OR pubmednotmedline[sb] OR indatareview[sb] OR pubstatusaheadofprint)) AND (comment[Filter] OR editorial[Filter] OR letter[Filter]))

**Medline**

MEDLINE (Ovid) contains peer-reviewed journal articles, focusing on biomedical sciences such as medicine, nursing, and pre-clinical sciences (Chambliss, 1991).

1. Prejudice/ or Racism/

2. (racism or racial-discriminat* or racial-prejudice or racist-event* or racist-episode* or racial-stereotype* or race-related-stress).tw,kf.

3. ((discriminat* or bias* or prejudic* or hostil* or harass* or bully* or cyberbull* or cyber-bull* or (unfair* adj1 treat*) or oppress*) adj3 (race or racial* or ethnic* or cultur* or religio* or migrant* or refugee* or asylum)).tw,kf.

4. (newborn* or new-born* or baby or babies or neonat* or neo-nat* or infan* or toddler* or pre-schooler* or preschooler* or kinder or kinders or kindergarten* or boy or boys or girl or girls or child or children or childhood or pediatric* or paediatric* or adolescen* or youth or youths or teen or teens or teenage* or school-age* or schoolage* or school-child* or schoolchild* or school-girl* or schoolgirl* or school-boy* or schoolboy* or young-person* or young-people).af.

5. Child Welfare/ or pediatric obesity/et, ep, pc

6. (Prejudice/ or *Racism/ or 2 or 3) and 5

7. obesity/et, ep, pc or body mass index/ or overweight/pc

8. Waist-Hip Ratio/

9. Blood Pressure/ or Biomarkers/

10. Hypertension/et, ep, pc

11. exp Cardiovascular Diseases/et, ep, pc

12. depression/et, ep, pc or anxiety/et, ep, pc

13. Mental Health/

14. Stress, Psychological/et, ep, pc

15. Sleep/

16. exp Sleep Wake Disorders/et, ep, pc

17. "Quality of Life"/

18. Resilience, Psychological/ or exp adaptation, psychological/

19. exp substance-related disorders/et, ep, pc or alcohol-related disorders/et, ep, pc

20. smoking/et, ep or exp tobacco smoking/et, ep, pc

21. Mental Disorders/et, ep, pc

22. Self Concept/

23. personal satisfaction/

24. exp suicide/et, ep, pc

25. conduct disorder/et, ep, pc or aggression/et, ep, pc

26. pregnancy outcome/

27. (health-care or healthcare or health-service* or clinic? or ill-health or wellbeing or well-being or disease* or illness* or bmi or body-mass-index or anthropometric* or WHR or waist-hip-ratio or hypertension or blood-pressure or cardiometabolic or cardio-metabolic or biomarker* or obese or obesity or overweight or depress* or anxiety or anxious* or mental-health or mental-disorder* or stress or distress* or suicid* or sleep or psychosis or tobacco or smoke* or smoking or drug? or alcohol* or substance-use or substance-related-disorder* or resilien* or self-esteem or self-worth or self-concept or quality-of-life or life-satisfaction or personal-satisfaction or conduct-disorder* or aggression).tw,kf.

28. ((social or behavio* or emotion* or developmental* or psychological* or learning*) adj3 (difficul* or problem* or delay* or adjust*)).tw,kf.

29. (((pregnancy or birth or gestation*) and (outcome* or preterm or pre-term or premature or small-for-gestational-age)) or low-birthweight or low-birth-weight).tw,kf.

30. 7 or 8 or 9 or 10 or 11 or 12 or 13 or 14 or 15 or 16 or 17 or 18 or 19 or 20 or 21 or 22 or 23 or 24 or 25 or 26

31. *obesity/et, ep, pc or *body mass index/ or *overweight/pc or *Waist-Hip Ratio/ or *Blood Pressure/ or *Biomarkers/ or *Hypertension/et, ep, pc or exp *Cardiovascular Diseases/et, ep, pc or (*depression/et, ep, pc or *anxiety/et, ep, pc) or *Mental Health/pc or *Stress, Psychological/et, ep, pc or *Sleep/ or exp *Sleep Wake Disorders/et, ep, pc or *"Quality of Life"/ or (*Resilience, Psychological/ or exp *adaptation, psychological/) or (exp *substance-related disorders/et, ep, pc or *alcohol-related disorders/et, ep, pc) or (*smoking/et, ep or exp *tobacco smoking/et, ep, pc) or *Mental Disorders/et, ep, pc or *Self Concept/ or *personal satisfaction/ or exp *suicide/et, ep, pc or (*conduct disorder/et, ep, pc or *aggression/et, ep, pc) or *pregnancy outcome/

32. (Prejudice/ or *Racism/ or 2 or 3) and (27 or 28 or 29 or 31) and 4

33. 6 or 32

34. limit 33 to (comment or editorial or letter)

35. 33 not 34

**PsycINFO**

PsycINFO contains a range of journal articles, books, book chapters, dissertations, and conference proceedings, all in the fields of psychology and related behavioural and social sciences (Burman, 2018).

1. exp prejudice/ or "race and ethnic discrimination"/ or exp "racial and ethnic attitudes"/ or racism/
2. (racism or racial-discriminat* or racial-prejudice or racist-event* or racist-episode* or racial-stereotype* or race-related-stress).ti,ab,id.
3. ((discriminat* or bias* or prejudic* or hostil* or harass* or bully* or cyberbull* or cyber-bull* or (unfair* adj1 treat*) or oppress*) adj3 (race or racial* or ethnic* or cultur* or religio* or migrant* or refugee* or asylum)).ti,ab,id.
4. (newborn* or new-born* or baby or babies or neonat* or neo-nat* or infan* or toddler* or pre-schooler* or preschooler* or kinder or kinders or kindergarten* or boy or boys or girl or girls or child or children or childhood or pediatric* or paediatric* or adolescen* or youth or youths or teen or teens or teenage* or school-age* or schoolage* or school-child* or schoolchild* or school-girl* or schoolgirl* or school-boy* or schoolboy* or young-person* or young-people).af.
5. child welfare/
6. (1 or 2 or 3) and 5
7. body mass index/ or obesity/ or exp overweight/
8. waist-hip-ratio.ti,ab,id.
9. exp blood pressure/ or biological markers/
10. exp hypertension/
11. exp cardiovascular disorders/
12. exp major depression/ or exp anxiety disorders/ or anxiety/
13. exp mental health/
14. psychological stress/
15. sleep/
16. exp sleep wake disorders/
17. "quality of life"/
18. "resilience (psychological)"/ or emotional adjustment/
19. exp "substance use disorder"/
20. exp tobacco smoking/
21. mental disorders/
22. self-concept/
23. satisfaction/
24. exp suicide/
25. conduct disorder/ or aggressiveness/ or aggressive behavior/
26. pregnancy outcomes/
27. (health-care or healthcare or health-service* or clinic? or ill-health or wellbeing or well-being or disease* or illness* or bmi or body-mass-index or anthropometric* or WHR or waist-hip-ratio or hypertension or blood-pressure or cardiometabolic or cardio-metabolic or biomarker* or obese or obesity or overweight or depress* or anxiety or anxious* or mental-health or mental-disorder* or stress or distress* or suicid* or sleep or psychosis or tobacco or smoke* or smoking or drug? or alcohol* or substance-use or substance-related-disorder* or resilien* or self-esteem or self-worth or self-concept or quality-of-life or life-satisfaction or personal-satisfaction or conduct-disorder* or aggression).ti,ab,id.
28. ((social or behavio* or emotion* or developmental* or psychological* or learning*) adj3 (difficul* or problem* or delay* or adjust*)).ti,ab,id.
29. (((pregnancy or birth or gestation*) and (outcome* or preterm or pre-term or premature or small-for-gestational-age)) or low-birthweight or low-birth-weight).ti,ab,id.
30. 7 or 8 or 9 or 10 or 11 or 12 or 13 or 14 or 15 or 16 or 17 or 18 or 19 or 20 or 21 or 22 or 23 or 24 or 25 or 26
31. *body mass index/ or *obesity/ or exp *overweight/ or waist-hip-ratio.ti,ab,id. or (exp *blood pressure/ or *biological markers/) or exp *hypertension/ or exp *cardiovascular disorders/ or (exp *major depression/ or exp *anxiety disorders/ or *anxiety/) or exp *mental health/ or *psychological stress/ or *sleep/ or exp *sleep wake disorders/ or *"quality of life"/ or (*"resilience (psychological)"/ or *emotional adjustment/) or exp *"substance use disorder"/ or exp *tobacco smoking/ or *mental disorders/ or *self-concept/ or *satisfaction/ or exp *suicide/ or (*conduct disorder/ or *aggressiveness/ or *aggressive behavior/) or *pregnancy outcomes/
32. (exp *prejudice/ or *"race and ethnic discrimination"/ or exp *"racial and ethnic attitudes"/ or *racism/ or 2 or 3) and (27 or 28 or 29 or 31) and 4
33. 6 or 32
34. limit 33 to ("column/opinion" or "comment/reply" or editorial or letter or review-book)
35. 33 not 34

**ERIC**

ERIC provides extensive access to educational-related literature, covering a range of scholarly literature and resources such as journal articles, conferences, and government documents (Wright & Pullen, 2007).

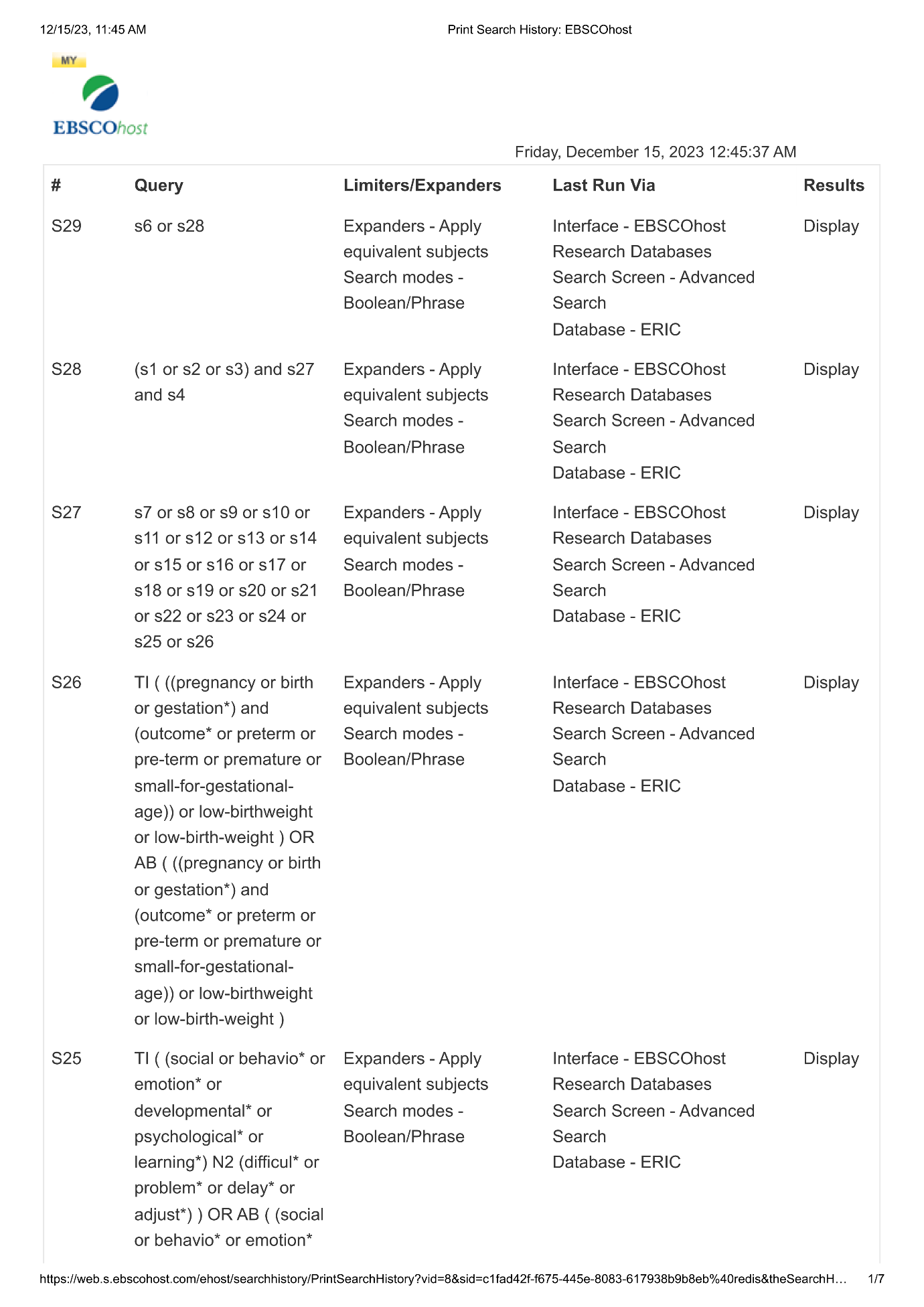

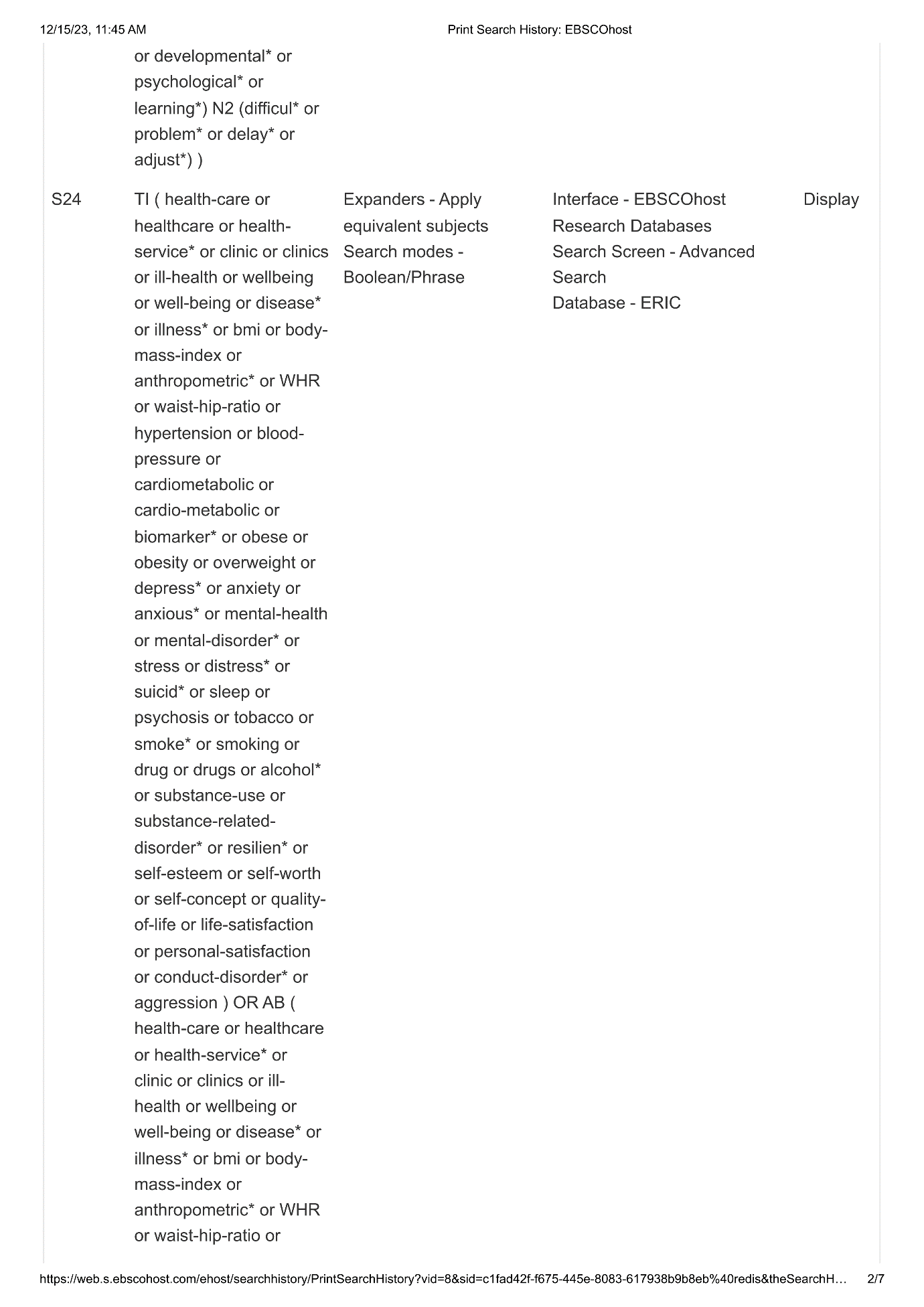

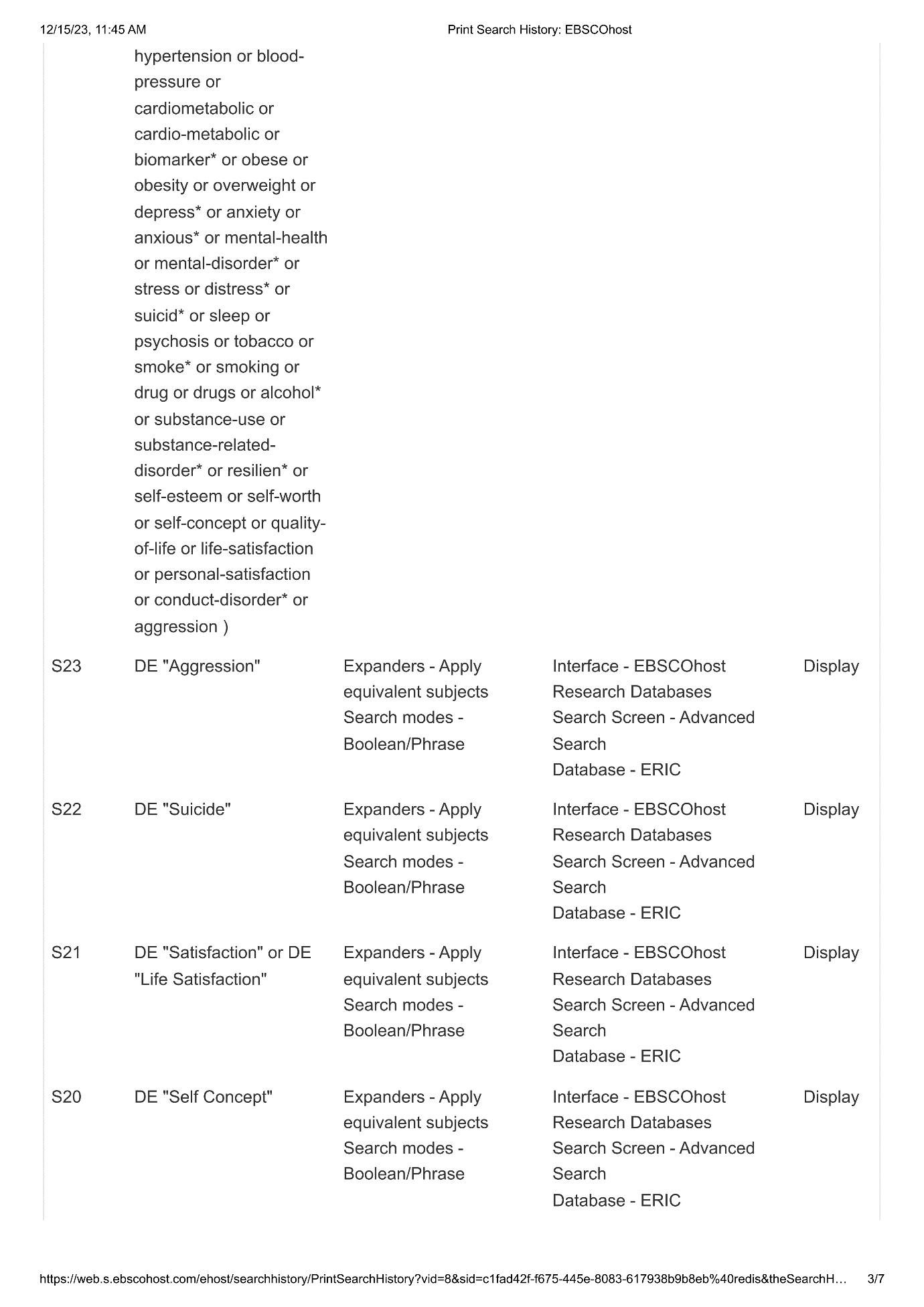

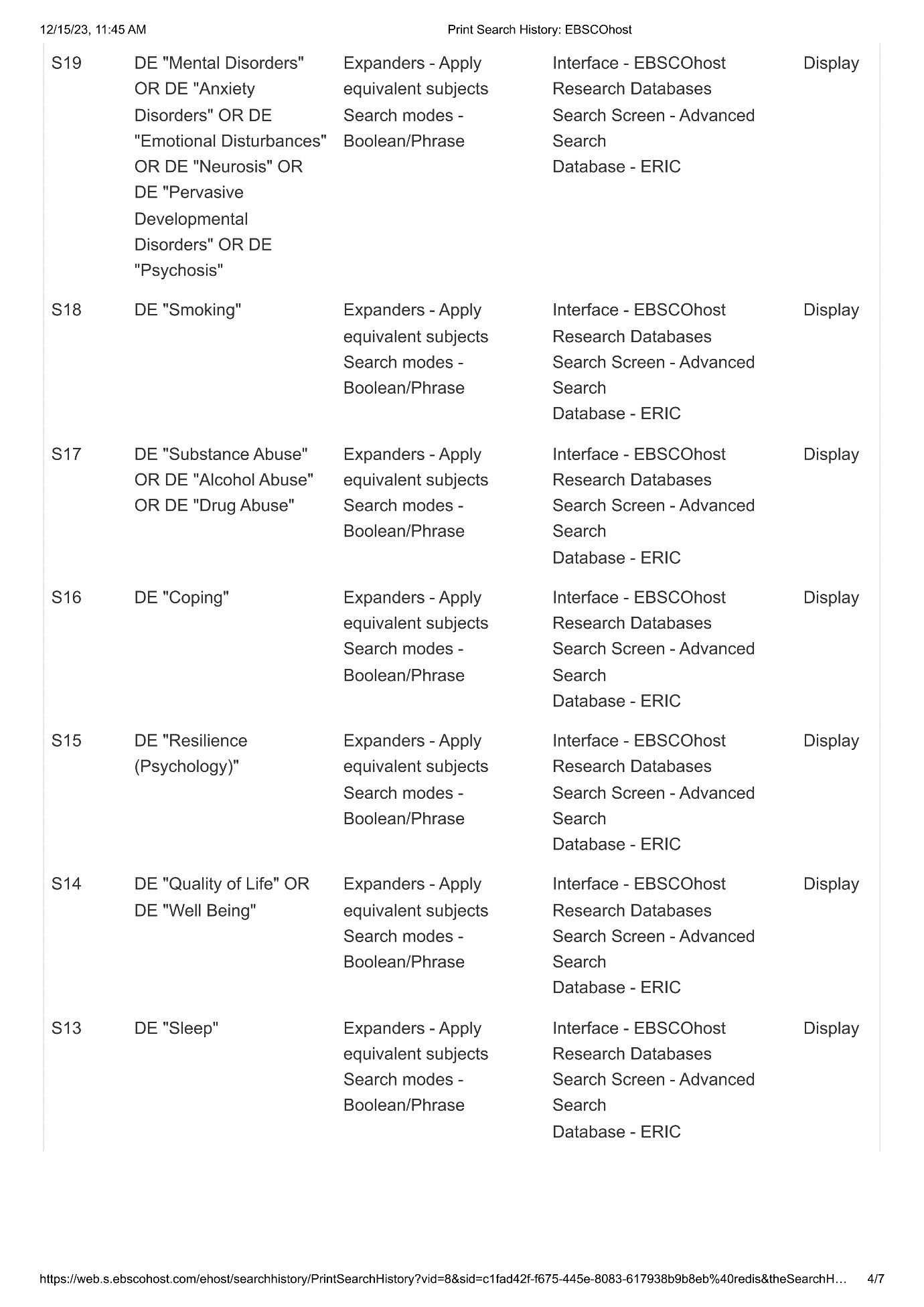

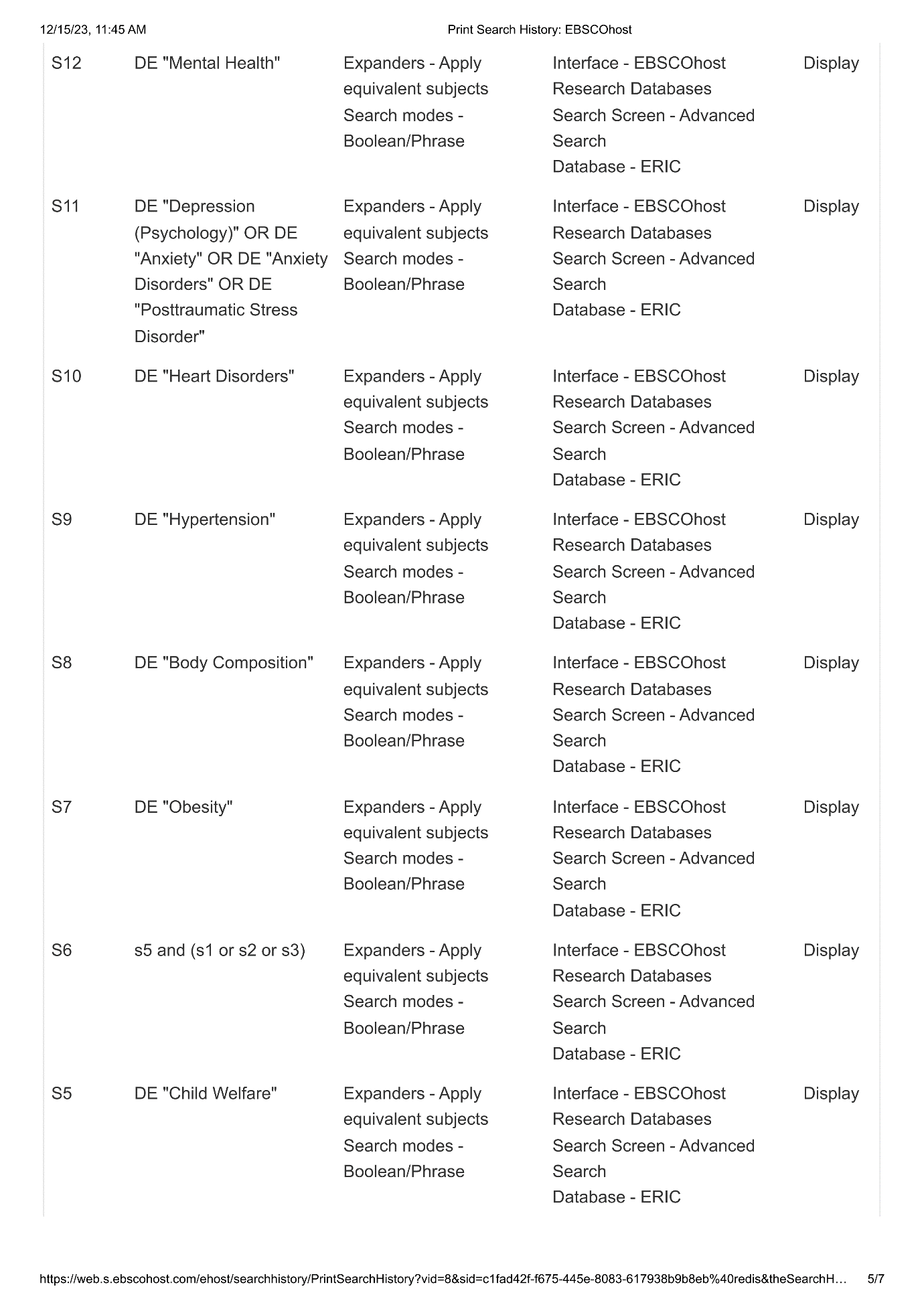

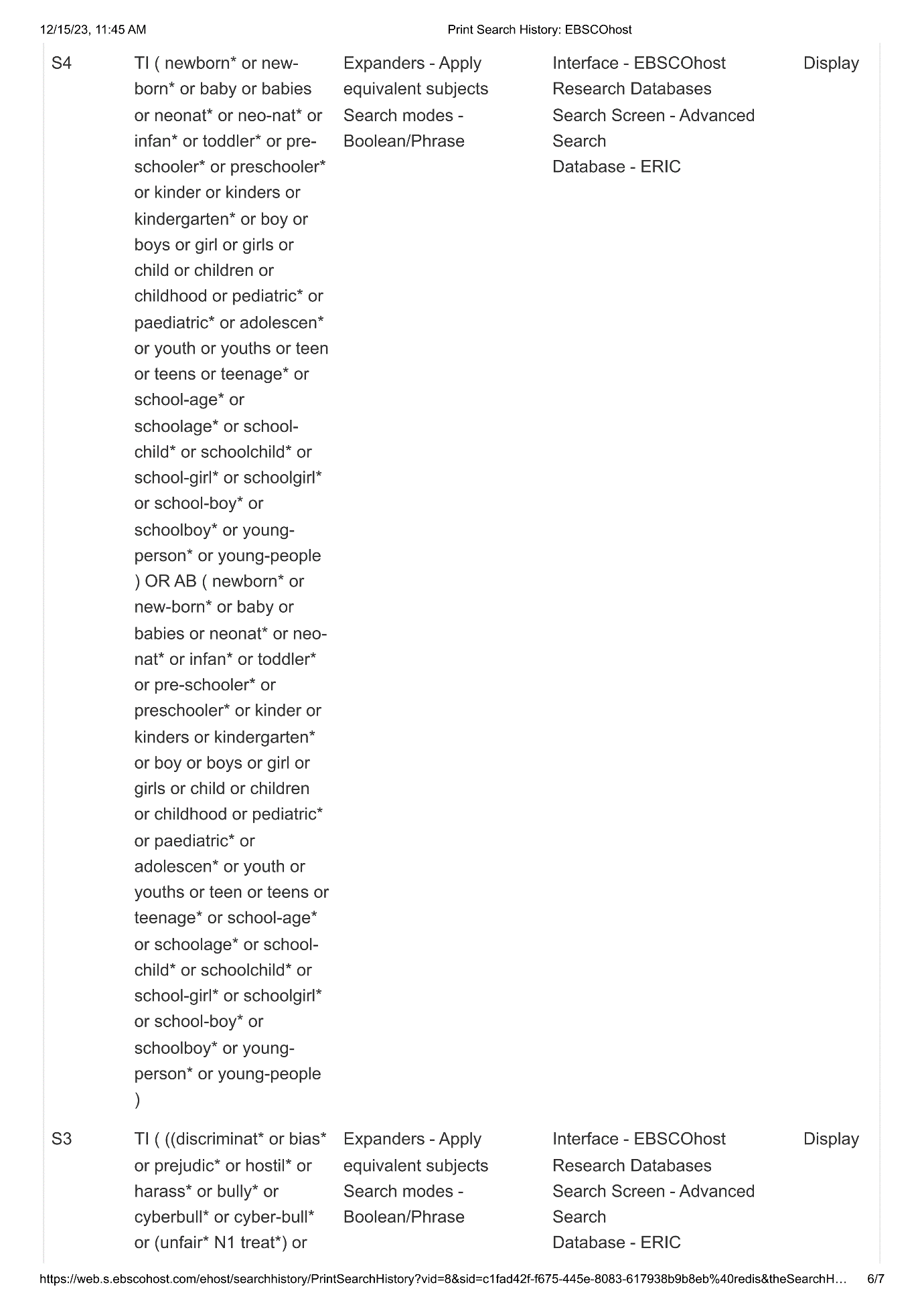

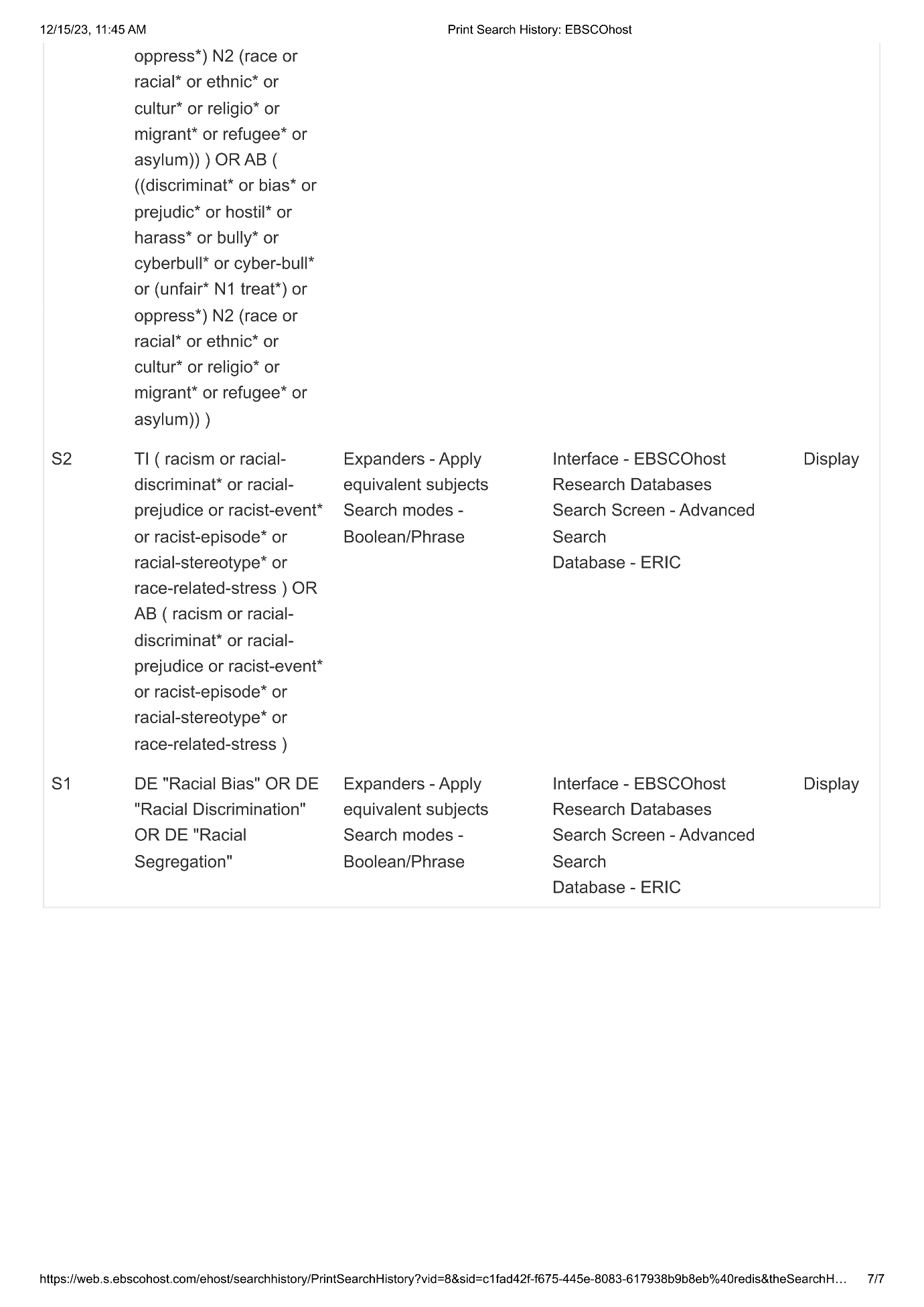
