## Supplementary material for "Racism and health and wellbeing among children and youth - an updated systematic review and meta-analysis": Supplementary file 5. Sensitivity analysis results.docx

**Supplementary file 6.** Sensitivity analysis results

Cortisol (Overall)

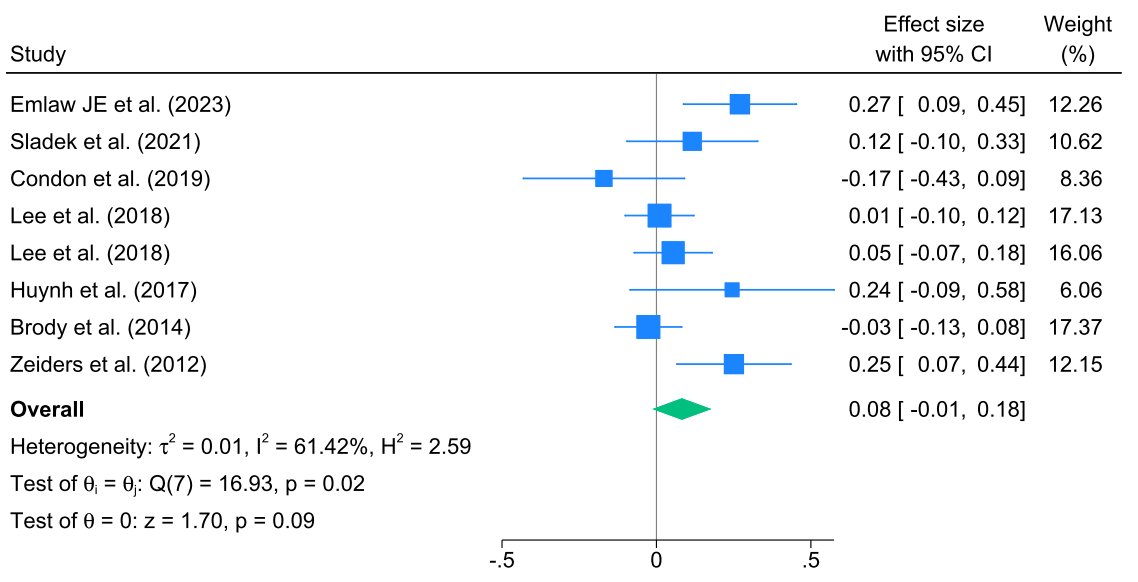

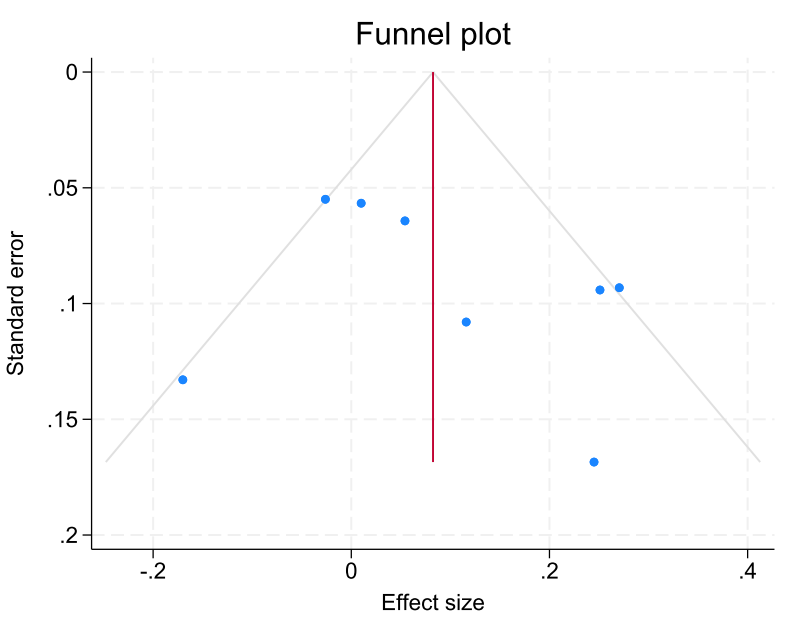

Supplementary Figure 7-1. Associations between racism and cortisol (overall) and the funnel plot.

Cortisol (saliva only)

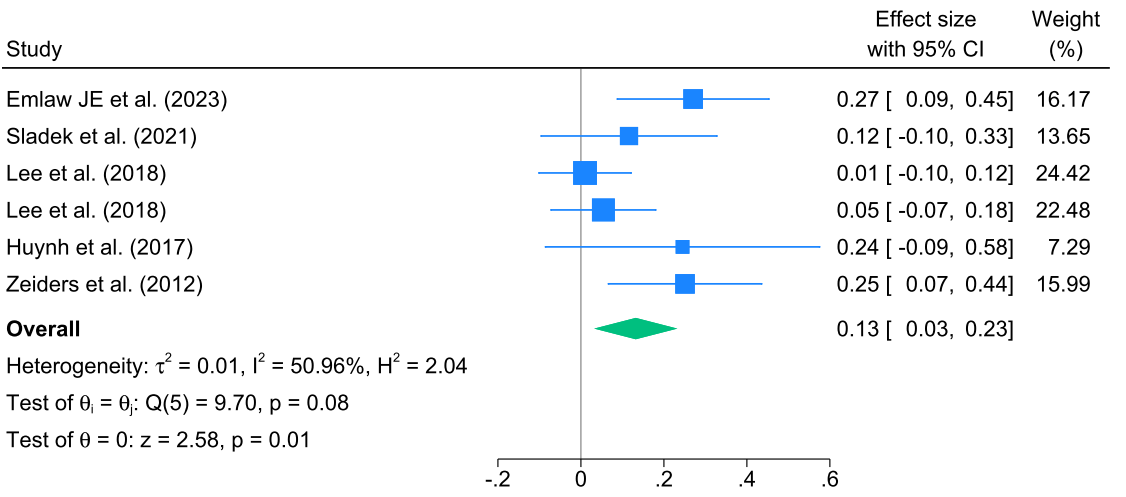

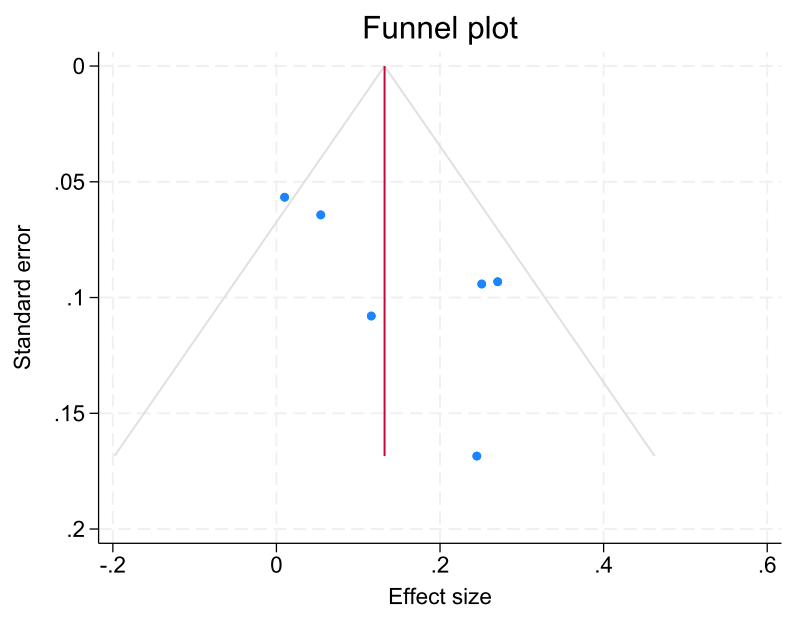

Supplementary Figure 7-2. Associations between racism and cortisol (saliva only) and the funnel plot.

C-reactive Protein (overall)

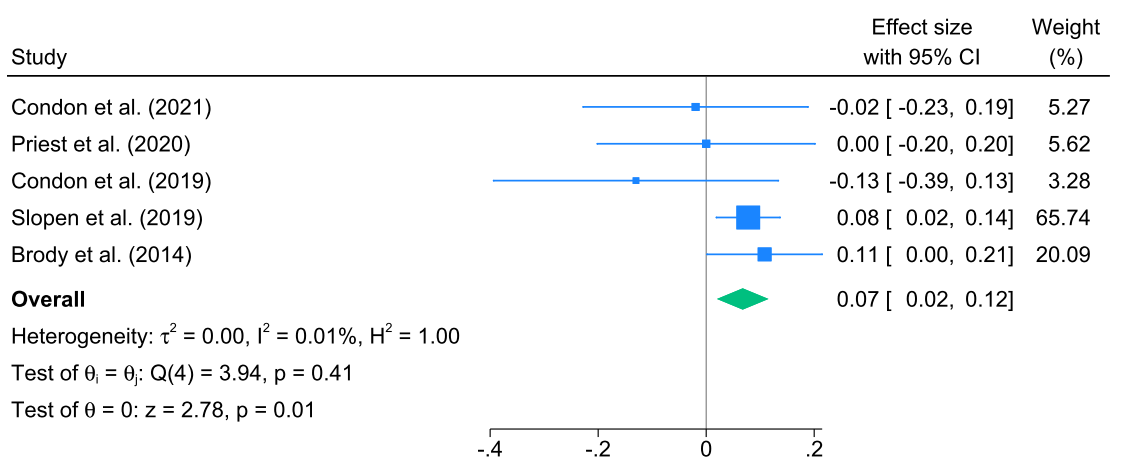

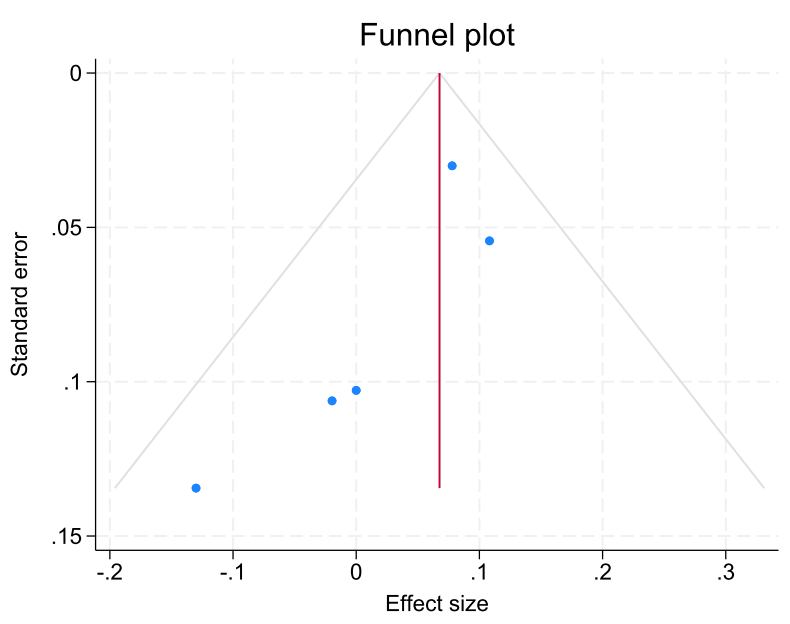

Supplementary Figure 8-1. Associations between racism and C-reactive Protein (overall) and the funnel plot.

C-reactive Protein (saliva only)

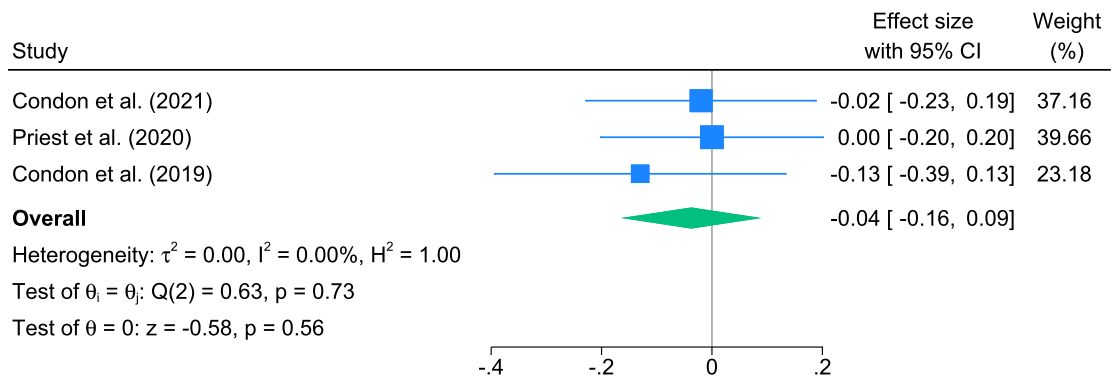

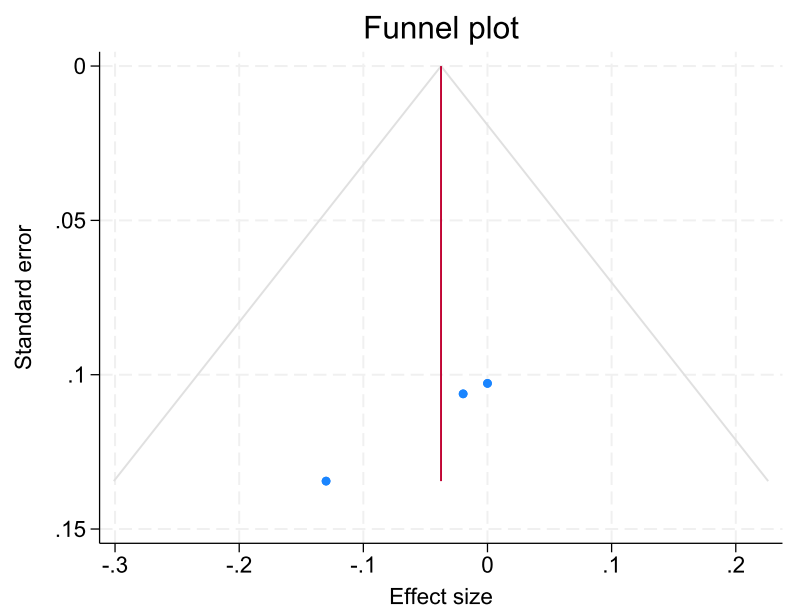

Supplementary Figure 8-2. Associations between racism and C-reactive Protein (saliva only) and the funnel plot.

IL-1β

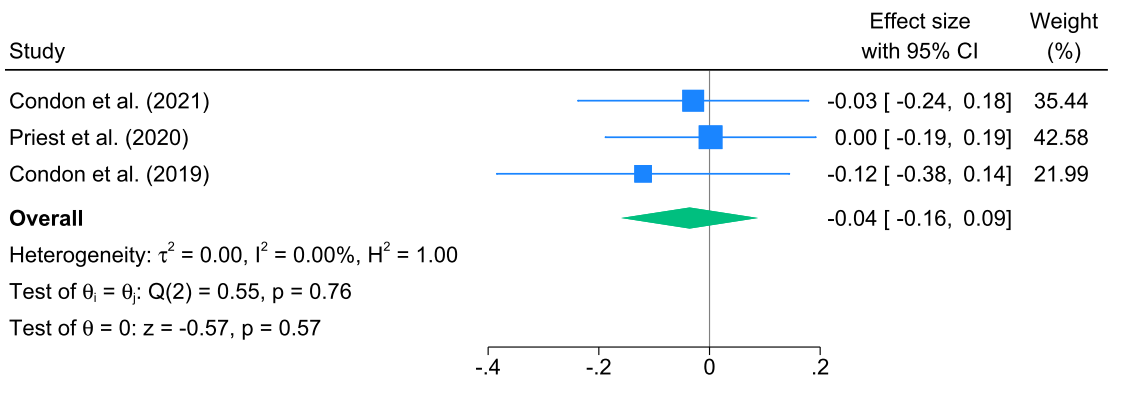

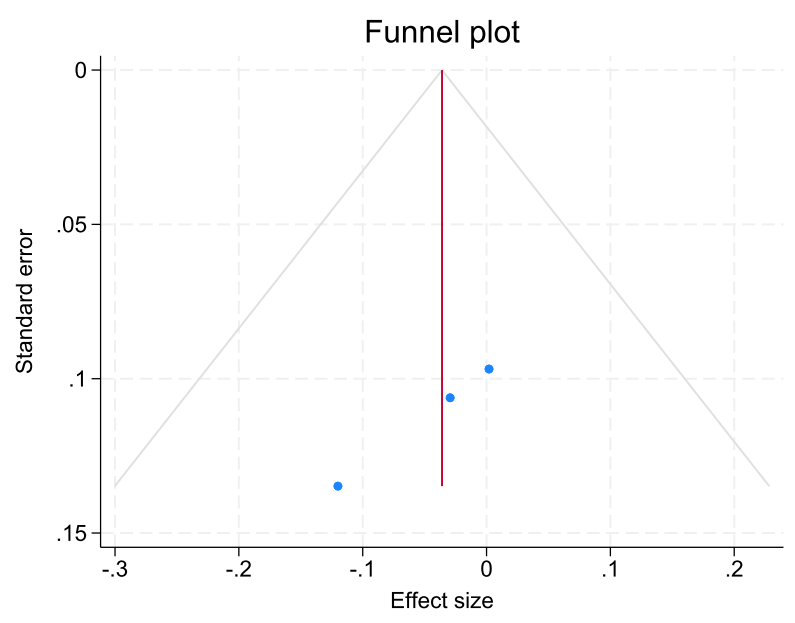

Supplementary Figure 9. Associations between racism and IL-1β and the funnel plot.

IL-6

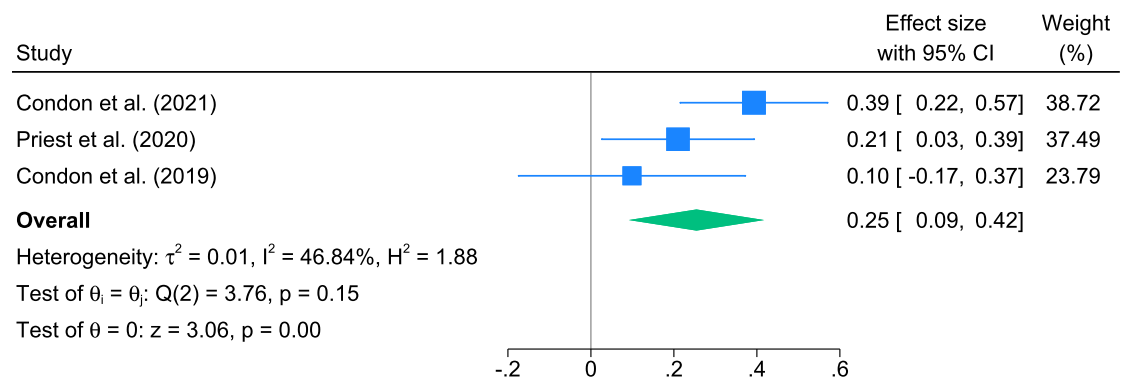

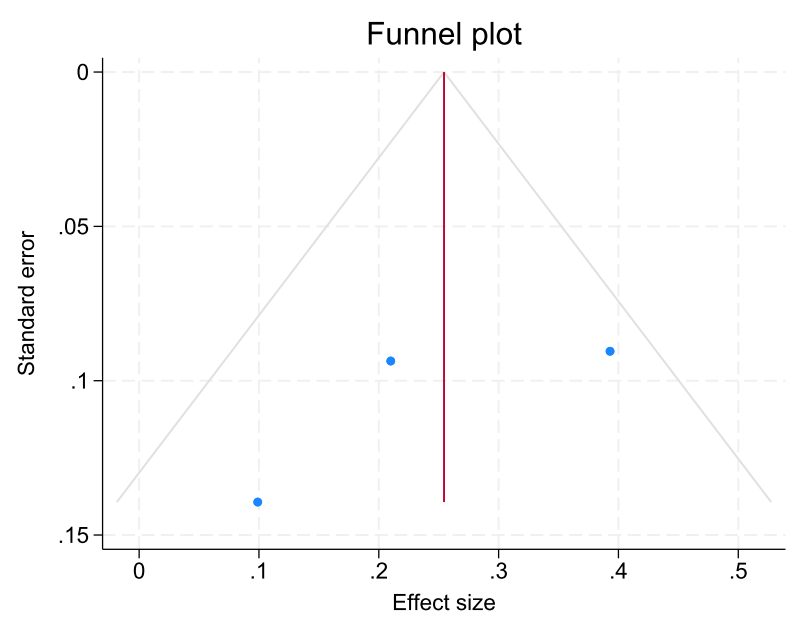

Supplementary Figure 10. Associations between racism and IL-6 and the funnel plot.

IL-8

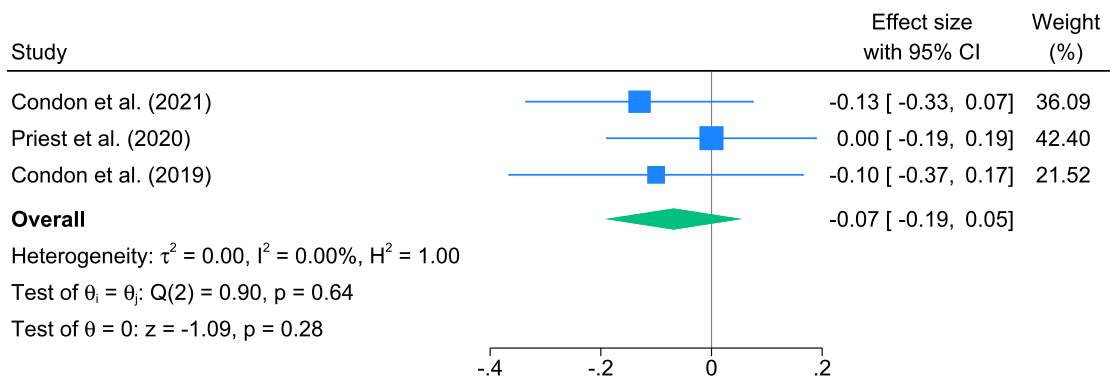

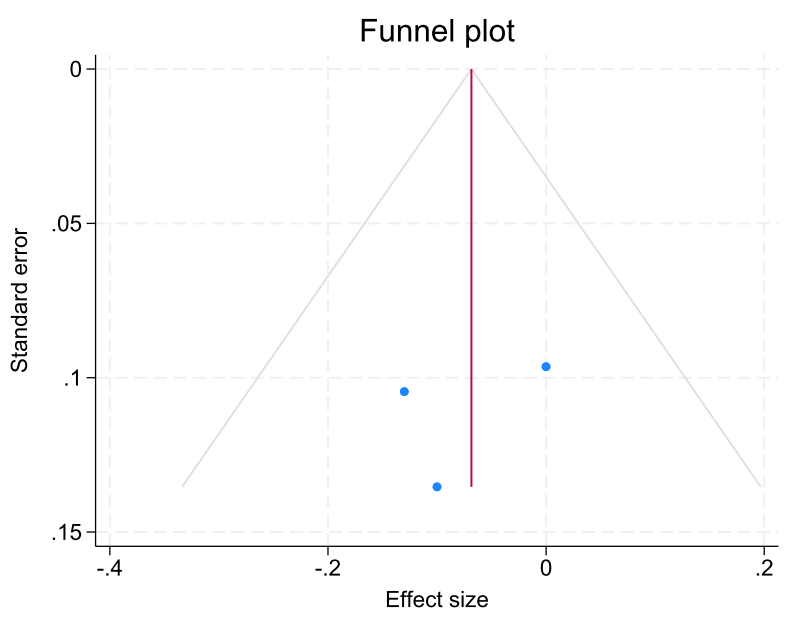

Supplementary Figure 11. Associations between racism and IL-8 and the funnel plot.

TNF- α

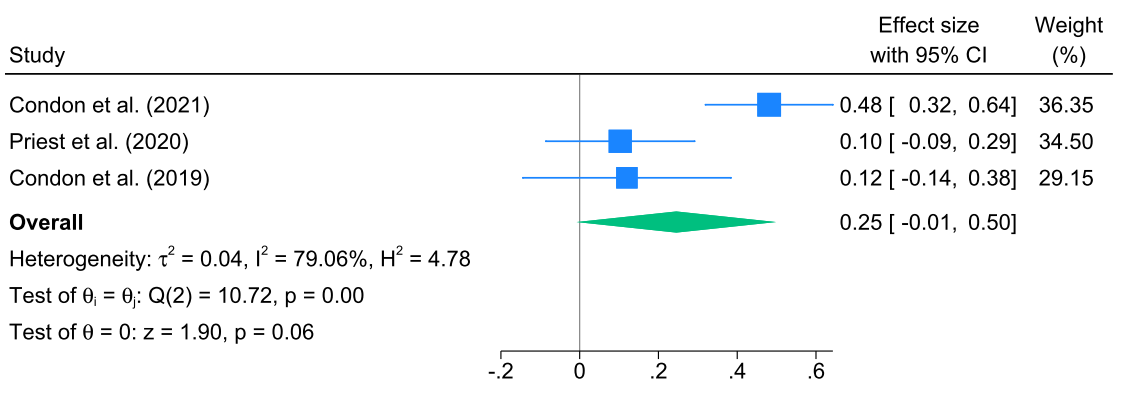

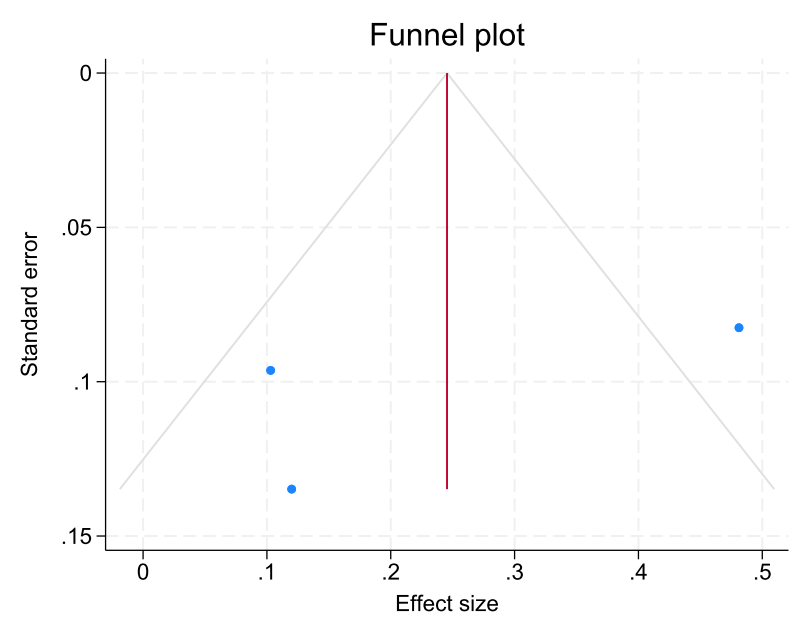

Supplementary Figure 12. Associations between racism and TNF- α and the funnel plot.

Asthma

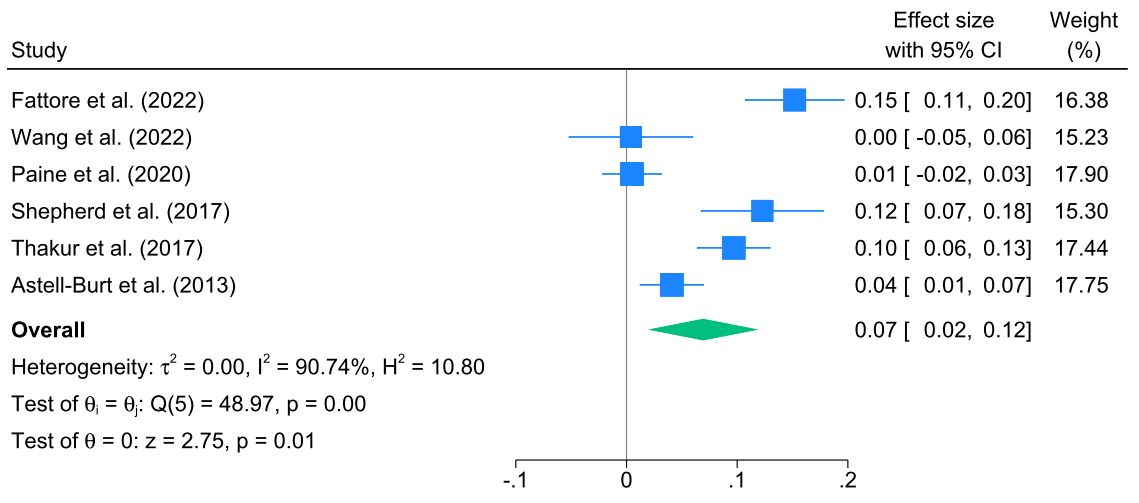

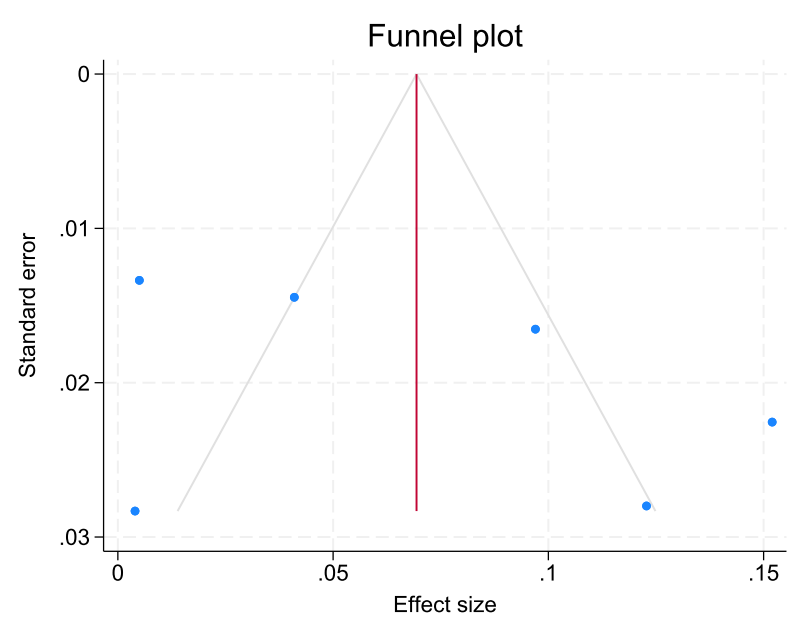

Supplementary Figure 13. Associations between racism and asthma and the funnel plot.

Somatic symptoms

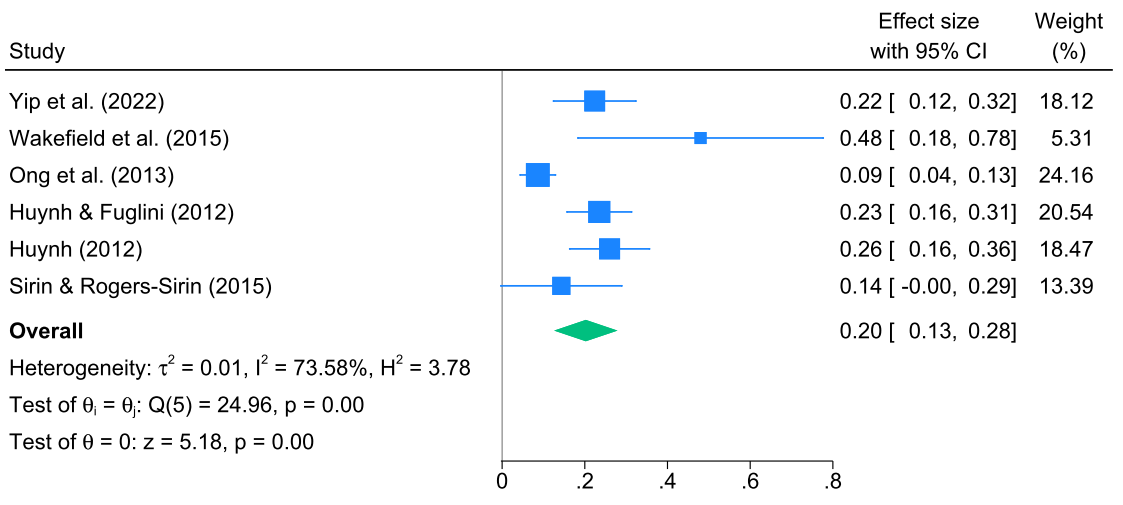

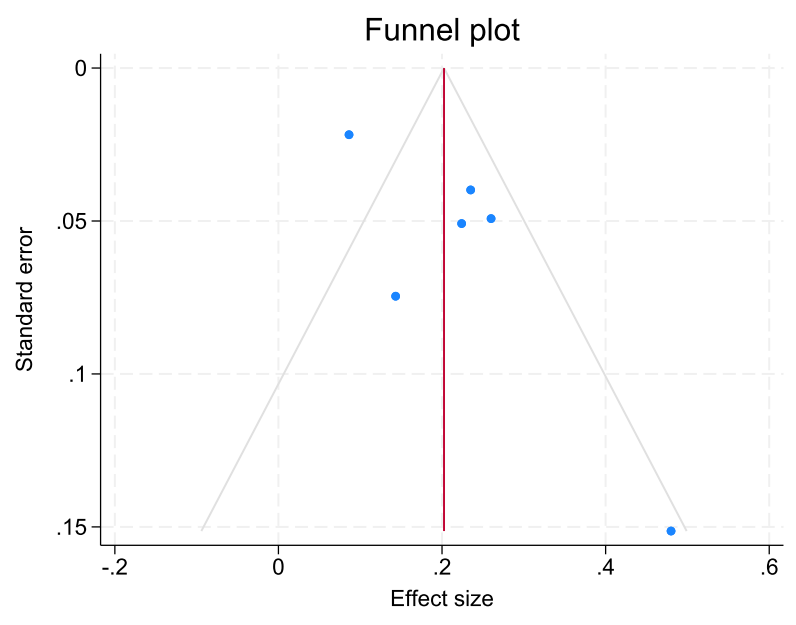
Supplementary Figure 14. Associations between racism and somatic symptoms and the funnel plot.

Systolic blood pressure

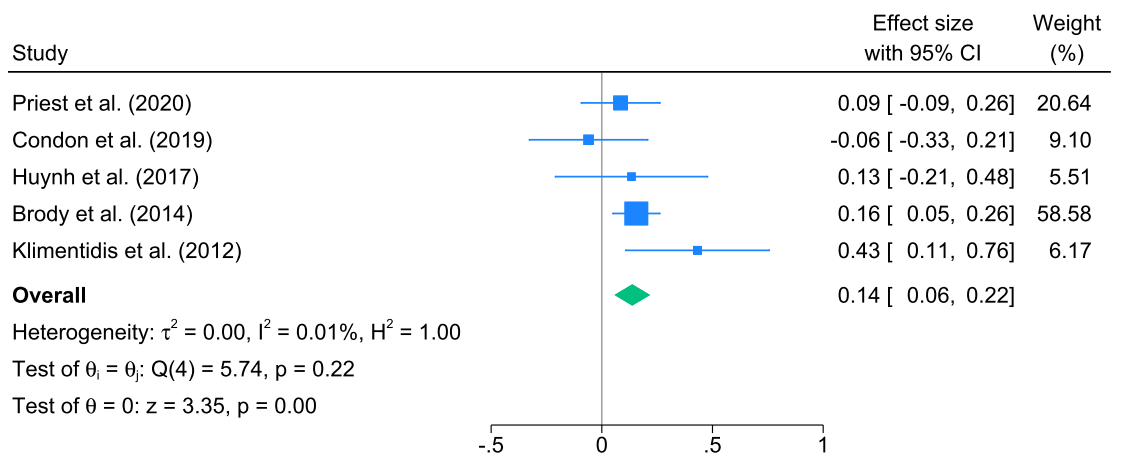

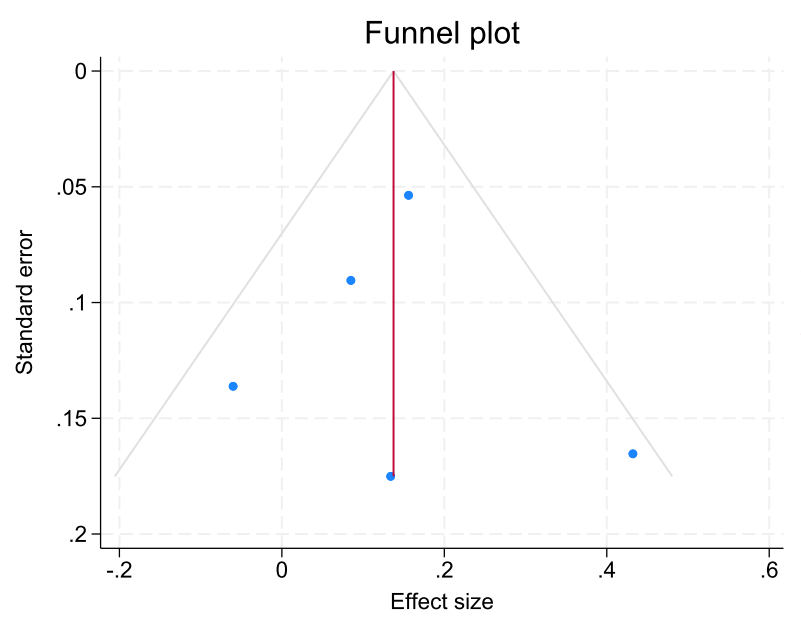

Supplementary Figure 15. Associations between racism and systolic blood pressure and the funnel plot.

Diastolic blood pressure

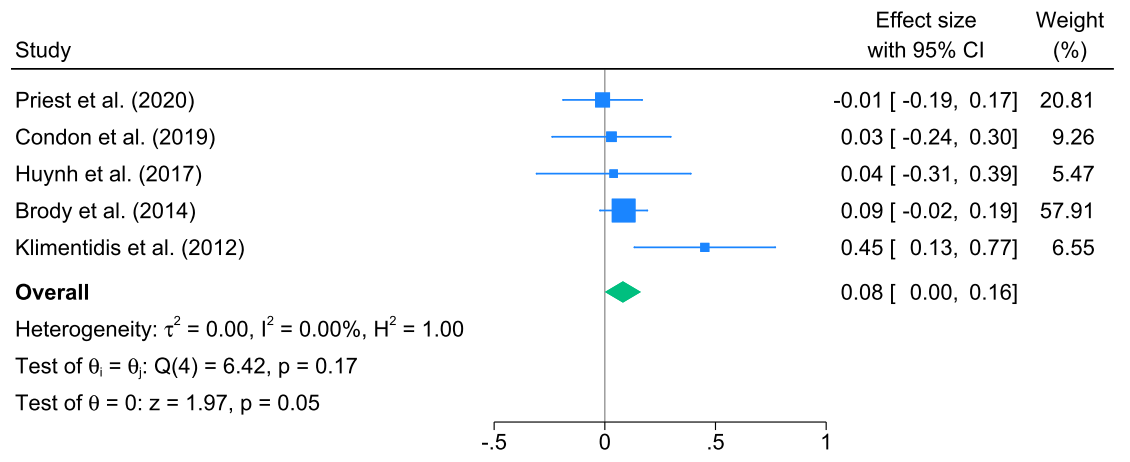

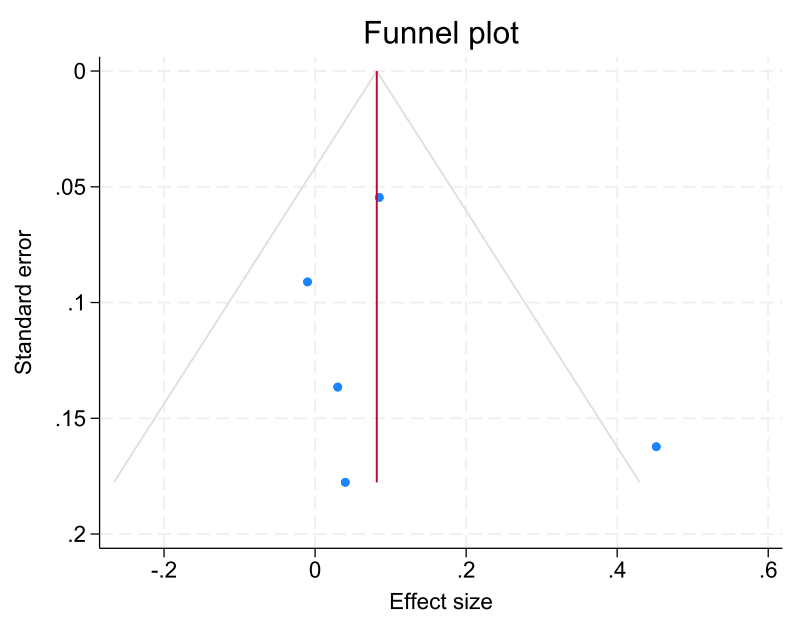

Supplementary Figure 16. Associations between racism and diastolic blood pressure and the funnel plot.

Body mass index (BMI) z-score

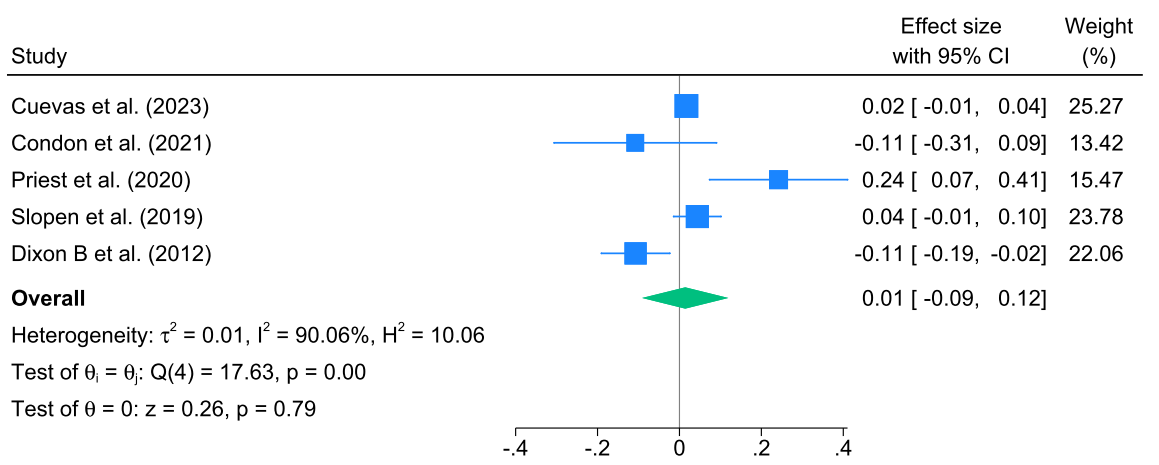

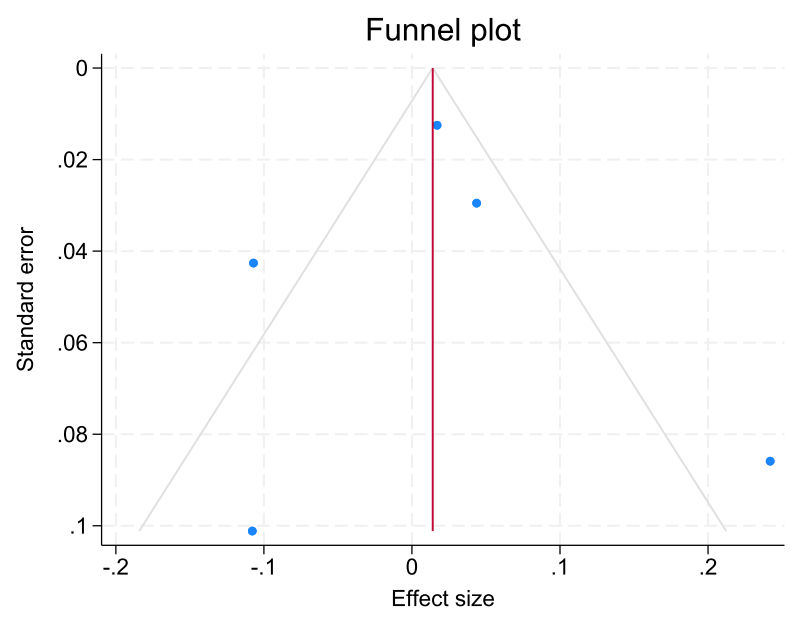

Supplementary Figure 17. Associations between racism and body mass index (BMI) z-score and the funnel plot.

BMI (kg/m^2^)

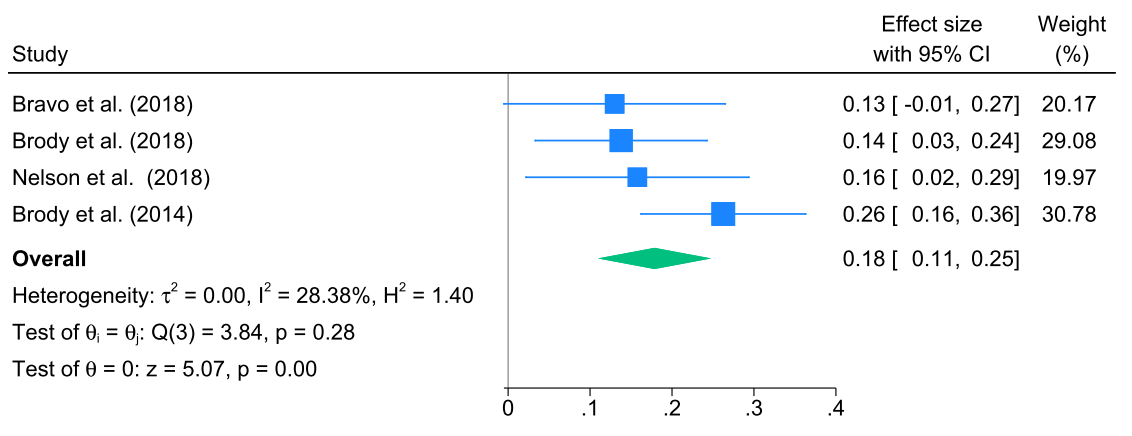

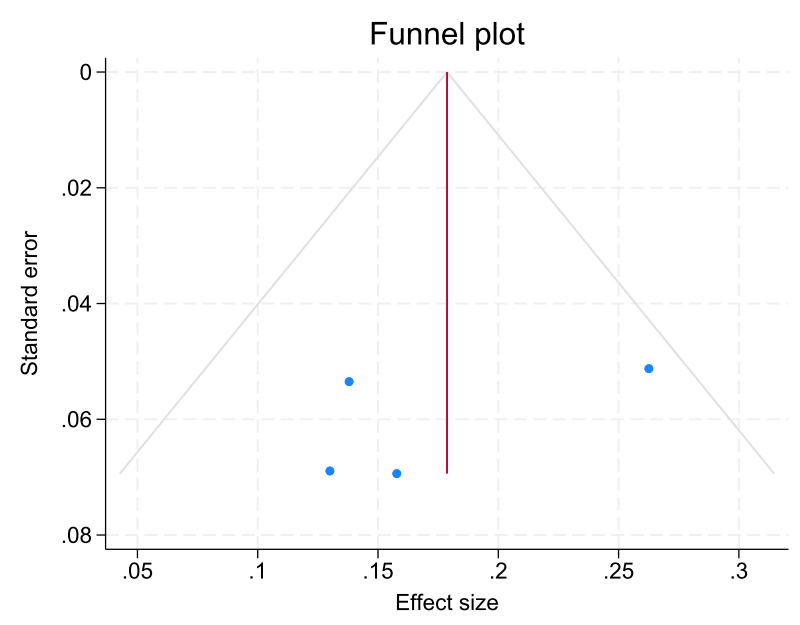

Supplementary Figure 18. Associations between racism and BMI (kg/m^2^) and the funnel plot.

Obesity

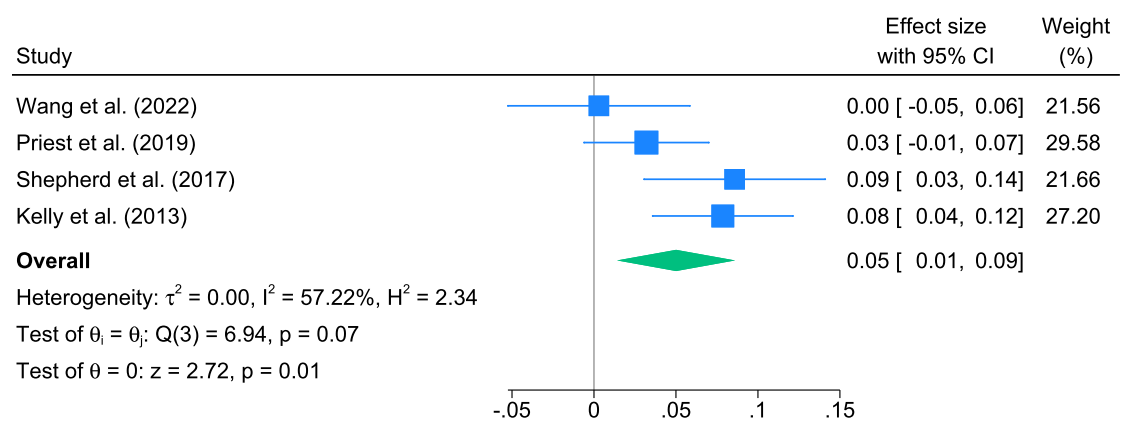

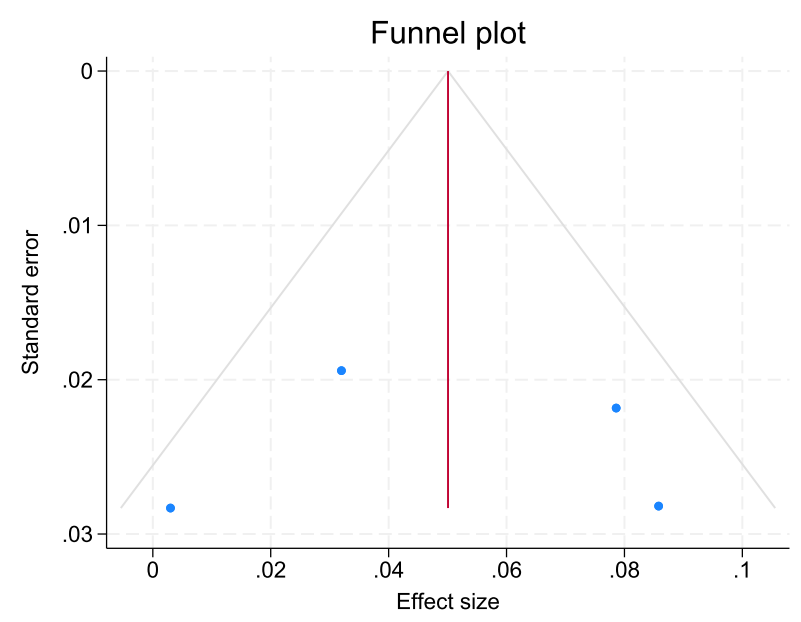

Supplementary Figure 19. Associations between racism and obesity and the funnel plot.
