## Supplementary material for "Racism and health and wellbeing among children and youth - an updated systematic review and meta-analysis": Supplementary file 6. Funnel plots results.docx

**Supplementary file 5. Funnel plots results**

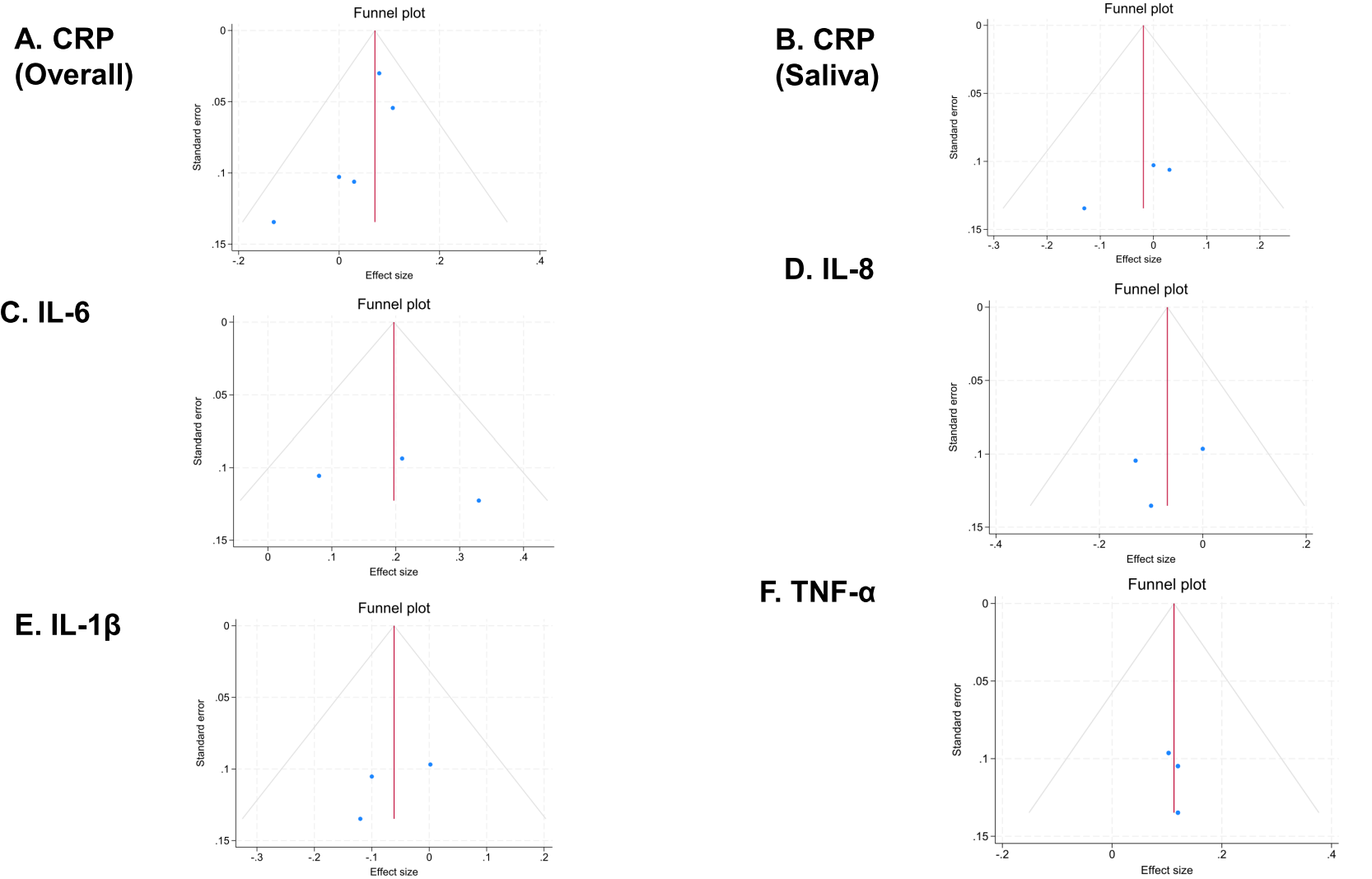

Supplementary Figure 1. Funnel plots for A) C-reactive Protein (overall); B) C-reactive Protein (saliva only); C) IL-6; D) IL-8; E) IL-1β; and F) TNF- α. Note: the red vertical line indicates the overall effect size.

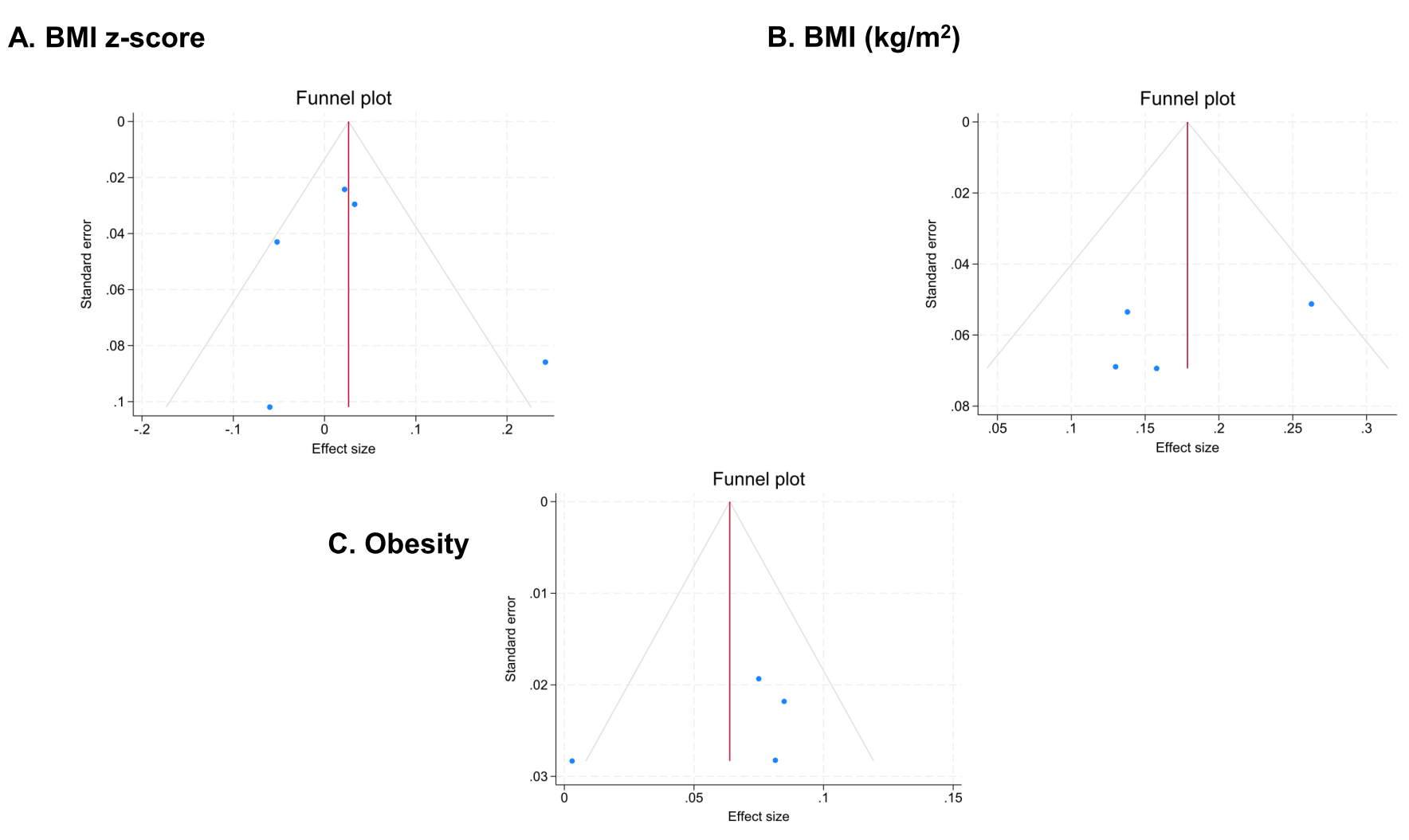

Supplementary Figure 2. Funnel plots for A) Body mass index (BMI) z-score; B) BMI; and C) Obesity. Note: the red vertical line indicates the overall effect size.

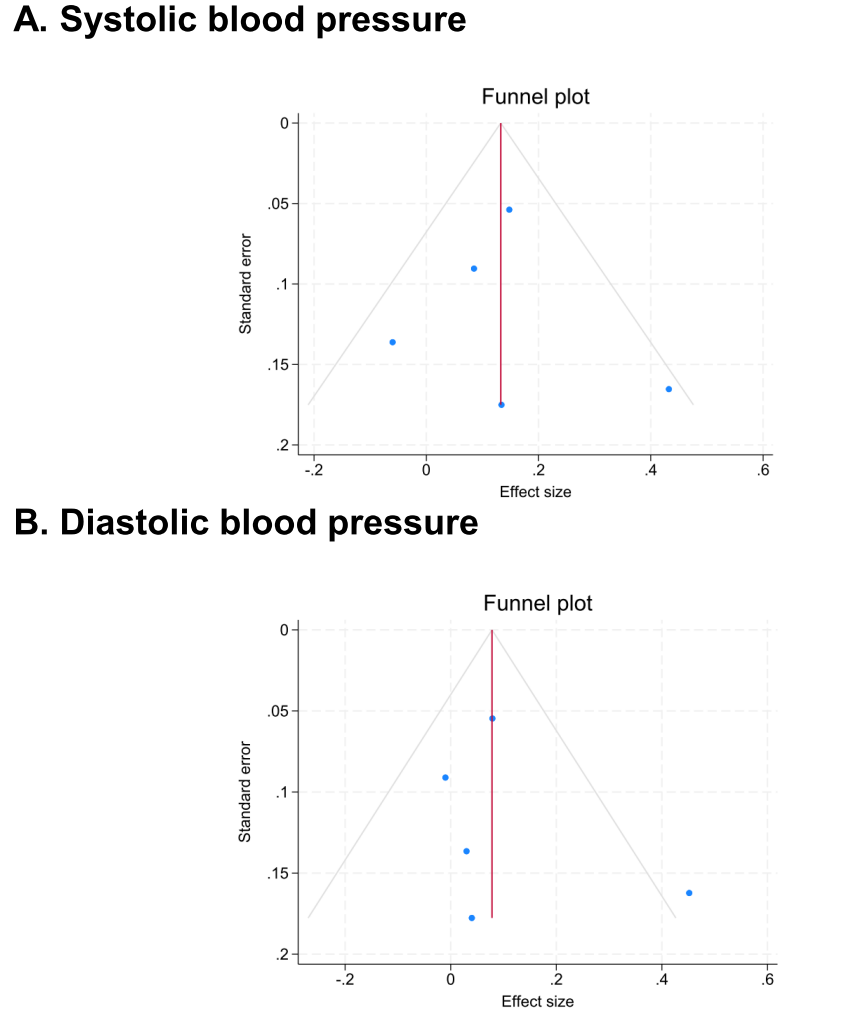

Supplementary Figure 3. Funnel plots for A) Systolic blood pressure; and B) Diastolic blood pressure. Note: the red vertical line indicates the overall effect size.

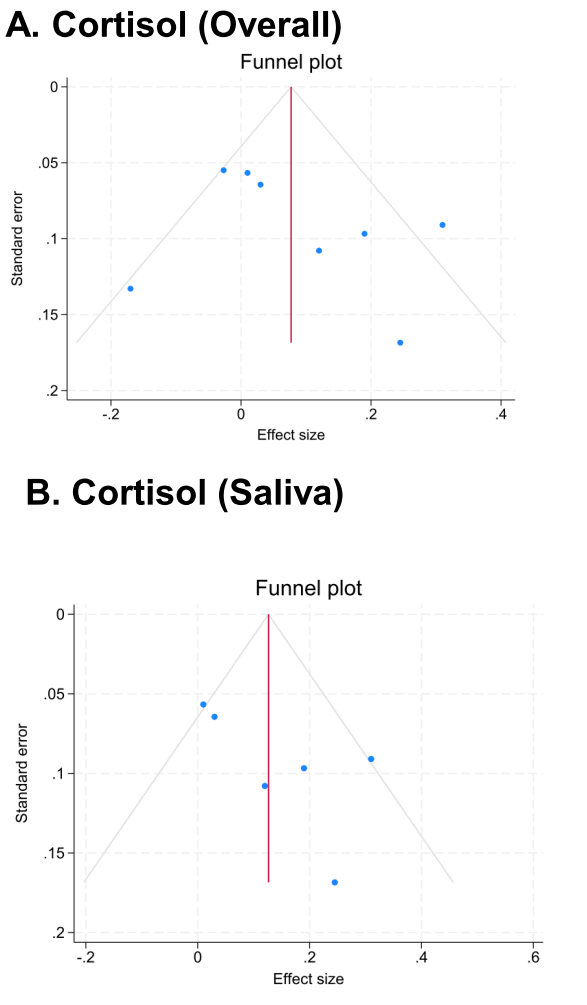

Supplementary Figure 4. Funnel plots for A) Cortisol (overall); and B) Cortisol (saliva only). Note: the red vertical line indicates the overall effect size.

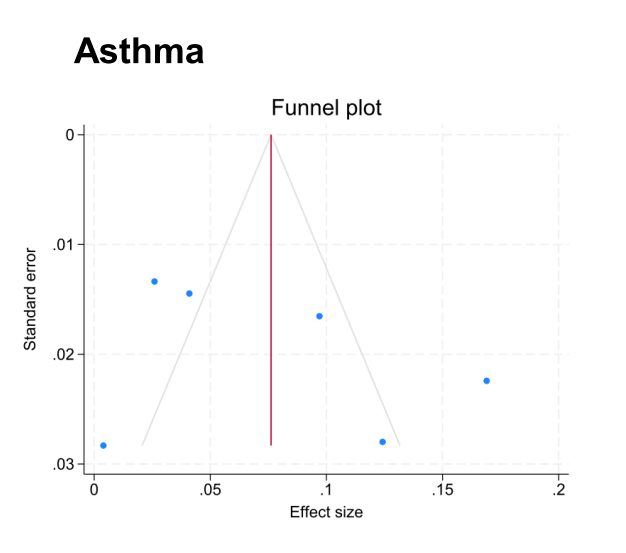

Supplementary Figure 5. Funnel plot for asthma. Note: the red vertical line indicates the overall effect size.

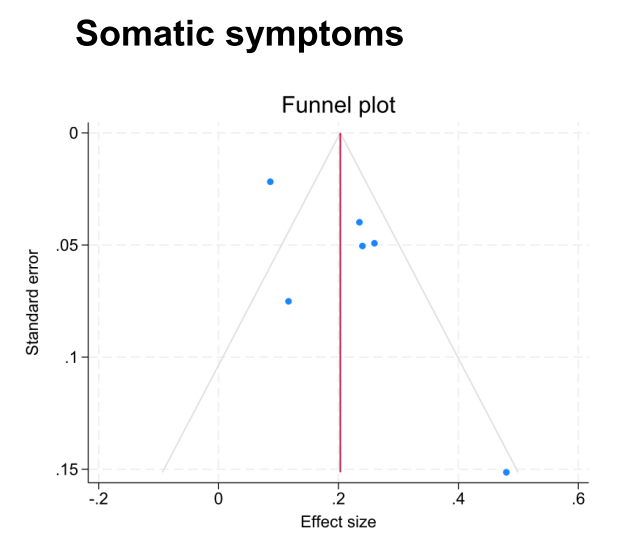

Supplementary Figure 6. Funnel plot for somatic symptoms. Note: the red vertical line indicates the overall effect size.
